# Missense and Loss of Function Variants at GWAS Loci in Familial Alzheimer’s Disease

**DOI:** 10.1101/2023.12.18.23300145

**Authors:** Tamil Iniyan Gunasekaran, Dolly Reyes-Dumeyer, Kelley M. Faber, Alison Goate, Brad Boeve, Carlos Cruchaga, Margaret Pericak-Vance, Jonathan L. Haines, Roger Rosenberg, Debby Tsuang, Diones Rivera Mejia, Martin Medrano, Rafael A. Lantigua, Robert A. Sweet, David A. Bennett, Robert S. Wilson, Camille Alba, Clifton Dalgard, Tatiana Foroud, Badri N. Vardarajan, Richard Mayeux

## Abstract

**BACKGROUND:** Few rare variants have been identified in genetic loci from genome wide association studies of Alzheimer’s disease (AD), limiting understanding of mechanisms and risk assessment, and genetic counseling.

**METHODS:** Using genome sequencing data from 197 families in The NIA Alzheimer’s Disease Family Based Study, and 214 Caribbean Hispanic families, we searched for rare coding variants within known GWAS loci from the largest published study.

**RESULTS:** Eighty-six rare missense or loss of function (LoF) variants completely segregated in 17.5% of families, but in 91 (22.1%) of families *APOE-e4* was the only variant segregating. However, in 60.3% of families neither *APOE-e4* nor missense or LoF variants were found within the GWAS loci.

**DISCUSSION:** Although *APOE-ε4* and several rare variants were found to segregate in both family datasets, many families had no variant accounting for their disease. This suggests that familial AD may be the result of unidentified rare variants.

## 1. BACKGROUND

In the United States (US) in 2023, Alzheimer’s disease (AD) was present in 6.7 million people, representing 10.8% of the population over age 65 years, and is projected to grow to 13.8 million people, nearly 3.3% of the total US population by 2060.^1^ The cause(s) of AD remain elusive, but there is strong evidence that it is a polygenic disorder. Highly penetrant pathogenic variants in AD: *APP*^2; 3^*PSEN1*^4^ and *PSEN2*^5^ explain less than 5% of early-onset familial AD (fAD). The *APOE-ε4* allele is associated with the risk of developing late onset, sporadic AD,^6; 7^and has been replicated in most ethnic groups.

Late-onset sporadic AD is highly heritable (∼70%), implying that genetic factors contribute to its etiology. The largest genome wide association studies (GWAS) that included thousands of unrelated individuals with AD and non-demented controls led by the European Alzheimer and Dementia Biobank Consortium, the UK Biobank, the Alzheimer’s Disease Genetic Consortium, the Alzheimer’s Disease Sequencing Project, FinnGen and CHARGE Consortia, identified common variants in more than 75 genetic risk loci. None of these risk loci reaches the effect size of *APOE-ε4*^8^.

The goal of this investigation was to identify rare variants associated within the 75 genetic loci in late-onset familial AD (fAD). We chose to limit the age at onset to later than 65 years to distinguish the study population from those with early-onset familial AD. This approach would allow us to estimate the proportion of families where the diagnosis of AD could be “explained” by genes identified by common variants in GWAS loci. Family-based statistical tests are largely underpowered to detect rare variants in novel genes with a limited number of family members and little or no evidence of vertical transmission. Thus, we used a simple segregation analysis restricted to genes that were previously associated with AD in GWAS.

There are clear advantages to family-based studies. Family-based analyses reduce the effects of population stratification, allow for easier detection of Mendelian inconsistencies and accurate haplotype phasing, and can detect the parent-of-origin effects. Investigation of well-characterized and carefully selected families can result in the identification of rare variants with significant roles in the pathogenesis of familial AD.^9^

One of the most frequent questions posed by family members of patients with fAD concerns familial risk. Polygenic risk scores have recently been used to assess recurrence risk and penetrance in families.^10^ However, identification of pathogenic variants in fAD, would advance mechanistic and therapeutic research efforts and facilitate genetic counseling. To identify and determine the frequency of potentially pathogenic variants in fAD, we used genome sequencing data from families recruited in the US and the Dominican Republic to identify damaging missense and loss of function (LoF) variants in candidate genes nominated from the largest GWAS analyses of clinically and pathologically diagnosed AD.^8^

## 2. METHODS

### 2.1 Inclusion Criteria and Ethical Review

We required first-degree family members, typically siblings, as the primary criterion^11^. The criteria for study entry included two living affected (clinically demented or cognitively symptomatic) siblings, 60 years of age or older, and a third living relative, similar in age, with or without dementia. We also included families with deceased affected siblings, as long as autopsy information or frozen brain tissue was available for affected members. Families in which participants were symptomatic, but did not meet criteria for AD, were still included with the commitment for follow-up. Details of the recruitment for both cohorts have been previously described^11–13^. Recruitment protocols underwent ethical review at each site and were consistent across all sites in the NIA AD-FBS. In the Dominican Republic there was a central review board for the EFIGA study following the review at Columbia University. Women and men were included. There were no exclusions based on ethnic group or race because this was a family-based study that investigated family-specific rare variants.

### 2.2 National Institute on Aging AD Family-Based Study (AD-FBS)

Beginning in 2002, families were recruited from Alzheimer’s Disease Centers across the United States. Inclusion criteria for this study required one member to have a diagnosis of definite or probable AD^14^ with onset after age 60 and a sibling with definite, probable, or possible AD and a similar age at onset. Another relative of the affected sibling pair, 60 years of age or older if unaffected, or if affected diagnosed with AD, was required. Any relatives over age 50 years were also recruited, regardless of cognitive status, but individuals deemed unaffected were required to have had documented cognitive testing and clinical examination. Most sites used a standard neuropsychological batter including Logical Memory (Immediate and Delayed), Selective Reminding Task, Orientation (MMSE), Digit Span Forward and Backward, Digit Ordering, Category Fluency (animals and vegetables)^11^. Every participant provided informed consent. Demographic information, previous clinical diagnoses, age at onset for patients with AD, method of diagnosis, Clinical Dementia Rating Scale^15^ was ascertained. The age at onset for participants was defined as the age at which the family first observed signs of impaired cognition. For unaffected family members, we used their age at the time of their latest examination without impairment. Participants with advanced disease or those living in a remote location that could not complete a detailed in-person evaluation, contributed a blood sample, and the site investigator conducted a detailed review of medical records or an assessment by telephone or videoconference to document the presence or absence of AD. We used the NINCDS-ADRDA criteria^14^ for definite AD or pathological criteria for AD^16; 17^ when based on postmortem information alone. Individuals with other forms of dementia, mild cognitive impairment or uncorroborated family reports of AD were considered as unaffected. From a pool of 251 families with 1,055 individuals, we restricted the selection to families with two or more affected family members by clinical or pathological diagnosis, which yielded 197 families with 926 individuals and WGS data. There were 545 (58.8%) individuals with fAD and 381 individuals without dementia. The mean age at onset was 75.2 (s.d.8.04) years of age and they were white, non-Hispanic. Those affected were significantly older than those unaffected (Table 1a). The majority of families were white, and of non-Hispanic ancestry. There were approximately 12 families of African American ancestry.

**Tables 1a and 1b.** Distribution of participating families, age at onset and APOE-ε4 frequency.

| **1a. NIA AD-FBS** | **Total families** | **Total participants** | **Affected** | **Unaffected** | **Age Affected** | **Age Unaffected** | **APOE-ε4 (%)** |
| --- | --- | --- | --- | --- | --- | --- | --- |
| Total Families | 251 | 1055 | 594 | 461 | 75.19±8.03 | 71.61±9.72 | 0.38 |
| Less than 2 affected | 54 | 79 | 60 | 19 | 76.18±6.17 | 81±2.94 | 0.42 |
| Greater than 2 affected | 197 | 926 | 545 | 381 | 75.21±8.04 | 70.56±9.41 | 0.38 |
| **1b. EFIGA** | **Total families** | **Total participants** | **Affected** | **Unaffected** | **Age Affected** | **Age Unaffected** | **APOE-ε4 (%)** |
| Total Families | 271 | 1480 | 833 | 647 | 71.97±9.85 | 66.54±8.77 | 0.33 |
| Less than 2 affected | 57 | 435 | 145 | 290 | 80.25±8.66 | 81.28±6.76 | 0.27 |
| Greater than 2 affected | 214 | 1340 | 793 | 547 | 71.91±9.88 | 66.11±8.72 | 0.33 |

### 2.3 Estudio Familiar de la Influencia Genética en Alzheimer (EFIGA) cohort

We acquired data from a second set of families with late onset fAD from the EFIGA cohort to identify variants segregating among families of Caribbean Hispanic ancestry. Study design, adjudication, and clinical assessment of AD in this cohort was previously described^12; 18^ and similar to that used in the AD-FBS. The neuropsychological batter for EFIGA included the Selective Reminding Test, Benton Recognition, Orientation (from MMSE), Abstract Reasoning (similarities and identities/oddities), Naming (Boston Naming Test), Verbal Fluency “CFL”, Category Fluency (animals), Repetition, Comprehension, Benton Matching, Attention, and Literacy^12^,^18^. All individuals provided informed consent. Participants were followed every two years and evaluated using the same neuropsychological battery, structured medical and neurological examination, assessment of depression,^19^ The Clinical Dementia Rating Scale,^15^ and the research clinical diagnosis of AD was determined. There were 271 families with 1,481 individuals but restricting the selection of families to those with two or more affected family members, 214 (78.9%) families remained with 1,340 individuals with available genome sequencing. There were 793 (59.2%) individuals who met criteria for clinical fAD and 547 individuals without dementia. The mean age of onset was 71.2 years (s.d.9.9). Those affected were significantly older that those unaffected (Table 1b). All of the individuals included with of Caribbean Hispanic ancestry.

### 2.4 Whole Genome Sequencing

The genome sequencing at the New York Genome Center and at the Uniformed Services University of the Health Sciences using one microgram of DNA, an Illumina PCR-free library protocol, and sequenced on the Illumina HiSeq platform. We assessed sex mismatches and unexpected familial relationships prior to analyses. We harmonized the WGS from the two sites in the EFIGA and AD-FBS families, and jointly called variants to create a uniform, analysis set. Genomes were sequenced to a mean coverage of 30x. Sequence data analysis was performed using an automated analysis pipeline that matched the CCDG and TOPMed recommended best practices.^20^ Briefly, sequencing reads were aligned to the human reference, hs38DH, using BWA-MEM v0.7.15. Variant calling was performed using the GATK best-practices. Variant filtration was performed using Variant Quality Score Recalibration (VQSR at tranche 99.6%) which identified annotation profiles of variants that could be called with high confidence and assigns a score (VQSLOD) to each variant. Variants passing the VQSR threshold were further filtered out for sample missingness (>2%), depth of coverage-DP<10 and genotype quality-GQ>20. We then annotated high quality variants using ANNOVAR. Specifically, variants were annotated for population level frequency using Genome Aggregation Database (gnomAD), *in-silico* function using Variant Effect Predictor (VEP) and variant conservation using Combined Annotation Dependent Depletion score (CADD).

### 2.5 Segregation analysis and identification of damaging missense and loss of function variants

The search was limited to variants in genetic loci identified in the largest GWAS where individuals were clinically or pathologically examined.^8^ This analysis included cohorts such as UK Biobank where AD diagnosis was ascertained by proxy or family history. We also searched genes involved in early onset fAD: *APP, PSEN1 and PSEN2*, and *AKAP9.* The total number of genes or loci queried was 81 (Supplementary Table 1). Segregation of variants was defined by presence in all affected family members and in an unaffected family member if they were at least five years younger than the mean age of onset of AD in the family. Damaging coding variants were prioritized: loss of function (LoF) variants (stoploss, stopgain, frameshift or splice variants) or damaging missense with a high CADD >20. Only variants considered rare in the general population (less that 1% in gnomAD) were included. The high allelic frequency of *APOE-ε4* in the general population and in AD, required examination of the co-segregation within families with and without a segregation of damaging missense or LoF variants. Linkage Disequilibrium (LD) computation: We computed LD between the statistically significant common variant at each loci reported ^8^ and segregating rare variant using PLINK. We report the r^2^ between the variants as a measure of LD.

## 3.0 RESULTS

There were a combined 522 families with 2,535 individuals sequenced in the two datasets. After excluding families with less than two affected family members there remained 411 families with 2,266 individuals in the two datasets. In the AD-FBS, 926 individuals (58.9%) met criteria for definite or probable fAD (19.6% vs 80.4%, respectively) and the remaining family members were unaffected. In EFIGA, there were 1,340 individuals in this family group, of which 59.1% met criteria for probable fAD and the remaining individuals were considered unaffected. Other demographics provide age at recruitment, age at onset and sex (Tables 1a & 1b).

### 3.1 Presence of damaging missense and LoF variants

We identified 4,749 total coding variants in 81 candidate genes and loci within the two cohorts (Supplementary table 1). We restricted the search to damaging potentially pathogenic coding variants using LoF variants or missense variants with CADD score greater than 20 (Figure 1). Forty-six damaging coding variants or loss of function variants were identified in 32 genes among 38 white, non-Hispanic families in AD-FBS. In the EFIGA there were 40 coding variants in 32 genes segregating in 34 families. *ABCA1, AKAP9, ANK3, NYO15A, and SLC26A1* contained the highest number of variants. No damaging missense variant was present in both family datasets, but a rare LoF frameshift variant in *ABCA7*, (Ch19:1044708, GGGGCACCTGGT:G) was present in the AD-FBS and EFIGA datasets. A full list of the coding variants in both family datasets are provided in Table 2. Variants in 20 genes were also found in the case-control series from the Alzheimer’s Disease Sequencing Project (Supplementary Table 2). Over 100 damaging missense or LoF variants in the AD-FBS and in the EFIGA families showed incomplete segregation in families (Supplementary Table 3).

**Figure 1.**
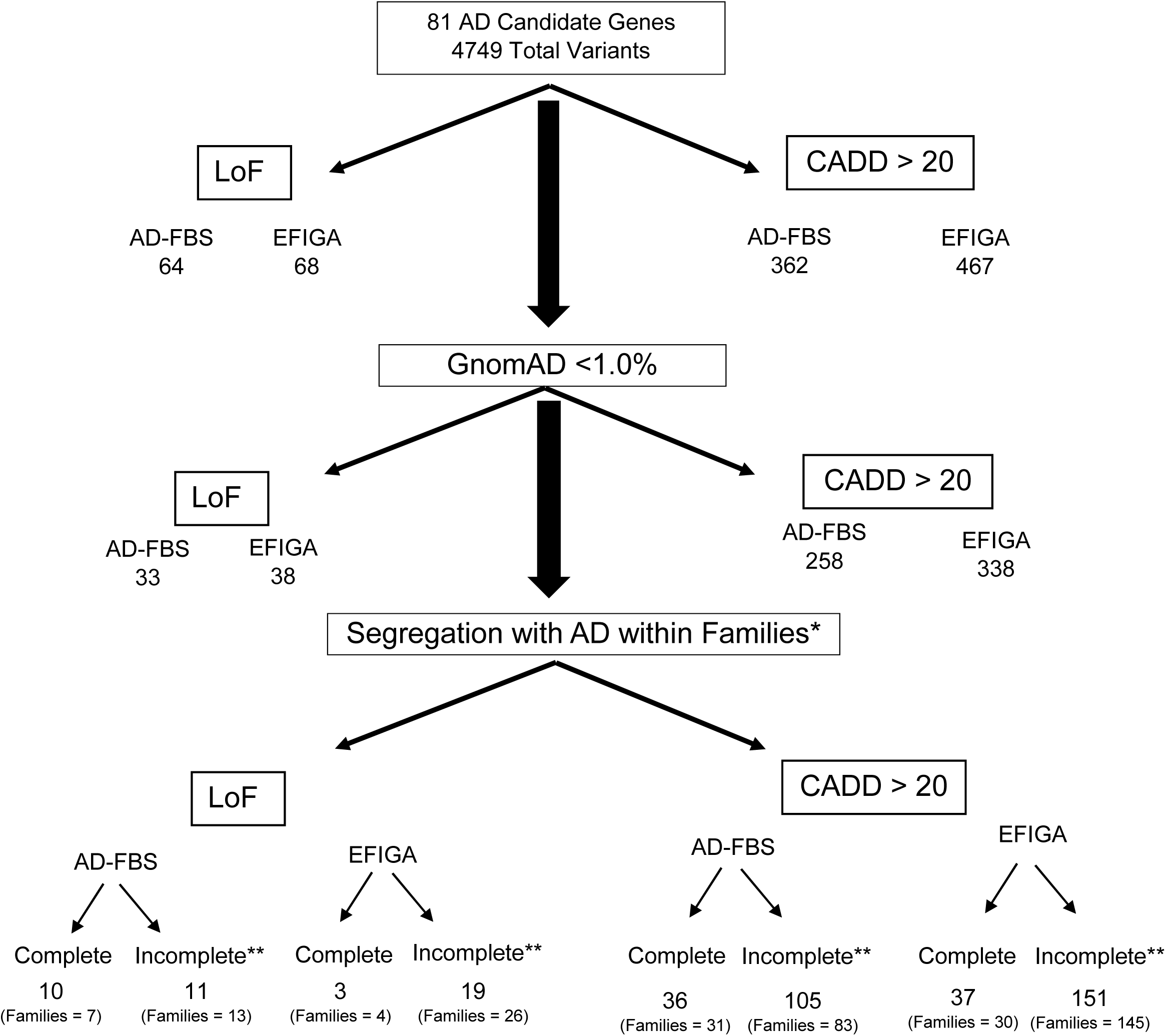
A filtering process was described in the methods. To identify rare variants (gnomAD less than 1%), degree of segregation (complete vs incomplete). Segregation was defined as being present in all affected members of a family. If a variant segregated in one family and showed incomplete segregation in another family, we required a qualifying variant to be seen only in affected. Unaffected family members with qualifying variants were accepted if their age was five or more years below the age at onset of the affected carriers. **Incomplete segregation was defined as variants present in at least four affected individuals with AD in the specific cohort (AD-FBS or EFIGA), observed in more affected than unaffected individuals, and if present in unaffected individuals if their age was five or more years below the age at onset of the affected carriers.

**Table 2.** Distribution of damaging missense and loss of function variants by cohort and variant type.

| Gene |  | Cohort | Variant Type | Reference |
| --- | --- | --- | --- | --- |
| *ABCA1* | chr9:104804721:C:G<br>chr9:104798567:T:C<br>chr9:104817343:G:A<br>chr9:104819624:C:T<br>chr9:104884475:G:A | AD-FBS<br>EFIGA<br>AD-FBS<br>AD-FBS<br>AD-FBS | LoF<br>Missense<br>Missense<br>Missense<br>Missense | Member of the superfamily of ATP-binding cassette transporters<sup>49</sup> |
| *ABCA7* | chr19:1057344:G:A<br>chr19:1051965:C:T<br>chr19:1044708:GGGGCACCTGGT:G | AD-FBS<br>EFIGA<br>EFIGA/<br>AD-FBS | Missense<br>Missense<br>LoF | Transmembrane transporter protein<sup>39; 40</sup> |
| *ABI3* | chr17:49217803:G:A | AD-FBS | Missense | Member of an adaptor protein family, also associated with Lewy Body disease<sup>50</sup> |
| *ACE* | chr17:63479054:G:C | EFIGA | Missense | Degradation and clearance of Aβ<sup>51</sup> |
| *ADAM10* | chr15:58717601:C:T | EFIGA | Missense | constitutive α-secretase processing of APP<sup>52</sup> |
| *ADAMTS1* | chr21:26838049:G:A<br>chr21:26838216:C:T | AD-FBS<br>EFIGA | Missense<br>Missense | APP hydrolysis decreased Aβ<sup>36</sup> |
| *ADAMTS20* | chr12:43356572:T:C<br>chr12:43428519:A:G | EFIGA<br>EFIGA | Missense<br>Missense | A disintegrin-like and metalloproteinase with thrombospondin type 1 motifs<sup>53</sup> |
| *AKAP9* | chr7:91992989:G:C chr7:92097077:C:A<br>chr7:92038422:A:G chr7:92040807:G:A<br>chr7:92102769:G:A | AD-FBS<br>AD-FBS<br>EFIGA<br>EFIGA<br>EFIGA | Missense<br>Missense<br>Missense | Enhances tau phosphorylation<sup>26</sup> |
| *ANK3* | chr10:60070700:T:A<br>chr10:60073965:T:C<br>chr10:60074069:A:C<br>chr10:60075299:G:A<br>chr10:60186803:G:A | EFIGA<br>EFIGA<br>AD-FBS<br>AD-FBS<br>AD-FBS | Missense<br>Missense<br>Missense<br>Missense<br>Missense | Alters methylation dynamics associated with aging specifically in neurons<sup>30</sup> |
| *BCKDK* | chr16:31110471:C:T | EFIGA | Missense | Associated with obesity and expression of AD related genes<sup>54</sup> |
| *CR1* | chr1:207511589:C:T<br>chr1:207523952:C:T | EFIGA<br>EFIGA | Missense | Immune dysfunction and glial-mediated neuroinflammation<sup>55</sup> |
| *CTSB* | chr8:11847108:T:G | AD-FBS | Missense | Accumulates in amyloid plaques in human AD brain<sup>56</sup> |
| *DOC2A* | chr16:30010237:T:C | AD-FBS | LoF | Microglial regulator of CNS innate immunity found in AD<sup>57</sup> |
| *ECHDC3* | chr10:11742730:C:T | EFIGA | LoF | Enoyl-CoA Hydratase Domain Containing 3 involved in lipid metabolism<sup>38</sup> |
| *EED* | chr11:86245326:C:T<br>chr11:86245267:C:T | EFIGA<br>AD-FBS | Missense<br>Missense | EED variants impact LTP, a measure of hippocampal memory function<sup>58</sup> |
| *EPHA1* | chr7:143398406:G:A<br>chr7:143391696:G:A | AD-FBS<br>EFIGA | Missense<br>Missense | Eph/ephrin signaling disrupts alters activity at the blood brain barrier <sup>59</sup> |
| *GPR141* | chr7:37741061:C:A<br>chr7:37741003:A:T | EFIGA<br>AD-FBS | Missense<br>Missense | Encodes an orphan receptor of the Class A rhodopsin-like G protein-coupled receptors<sup>60</sup> |
| *HS3ST5* | chr6:114057958:G:A | AD-FBS | Missense | Enhances the spread of tau pathology<sup>61</sup> |
| *IDUA* | chr4:987896:C:G | EFIGA/<br>AD-FBS | Missense<br>Missense | Accumulation of glycosaminoglycans<sup>62</sup> |
| *IL34* | chr16:70654657:C:A | AD-FBS | LoF | Physiological role in clearance of cerebral amyloid β-protein<sup>63</sup> |
| *LILRB2* | chr19:54279530:T:A | EFIGA | Missense | LILRB2 antagonist antibody on microglial responses to amyloid plaques<sup>64</sup> |
| *MYO15A* | chr17:18120185:G:A<br>chr17:18173824:G:A<br>chr17:18119509:G:A<br>chr17:18120434:C:T<br>chr17:18133274:G:A<br>chr17:18142122:G:A | EFIGA<br>EFIGA<br>AD-FBS<br>AD-FBS<br>AD-FBS<br>AD-FBS | Missense<br>Missense<br>Missense<br>Missense<br>Missense<br>Missense | Endosomal compartment disfunction<sup>65</sup> |
| *NME8* | chr7:37884404:G:T<br>chr7:37896956:C:CA | EFIGA<br>AD-FBS | LoF | Protective and antioxidant action<sup>66</sup> |
| *NYAP1* | chr7:100490675:C:T<br>chr7:100488976:C:T | AD-FBS<br>EFIGA | Missense<br>Missense | Affects PILRB and PILRA immunoglobulin-like receptors<sup>40</sup> |
| *PLCG2* | chr16:81858323:C:T<br>chr16:81923501:A:G | EFIGA<br>AD-FBS | Missense<br>Missense | Inflammatory response that is induced by amyloid plaques<sup>67</sup> |
| *PRDM7* | chr16:90060590:C:G<br>chr16:90062060:A:G | AD-FBS | Missense | Transcription and other nuclear processes<sup>68</sup> |
| *PRKD3* | chr2:37289451:T:TG<br>chr2:37289452:TACTC:T<br>chr2:37289460:T:TTG | AD-FBS | LoF | Inflammatory signaling<sup>69</sup> |
| *PSEN1* | chr14:73170945:C:T<br>chr14:73170959:A:G | AD-FBS<br>EFIGA | Missense<br>Missense | Subunit of γ-secretase <sup>70</sup> |
| *PTK2B* | chr8:27437198:G:A | EFIGA | Missense | Pyk2 suppresses Tau phosphorylation<sup>71</sup> |
| *RBCK1* | chr20:427337:C:T | AD-FBS | LoF | Heterologous ubiquitination<sup>72</sup> |
| *SFRP4* | chr7:37916317:C:T | EFIGA | Missense | Oxidative stress and inflammation<sup>73</sup> |
| *SLC26A1* | chr4:991652:G:A<br>chr4:989768:G:A<br>chr4:989896:A:G<br>chr4:989305:G:A<br>chr4:990080:G:A<br>chr4:990241:C:T | AD-FBS<br>EFIGA<br>EFIGA<br>AD-FBS<br>AD-FBS<br>AD-FBS | LoF<br>Missense<br>Missense<br>Missense<br>Missense<br>Missense | Member sulfate/anion transporter gene family<sup>32</sup> |
| *SNX1* | chr15:64126203:C:T<br>chr15:64131812:C:T | AD-FBS<br>AD-FBS | Missense<br>Missense | A member of the retromer complex<sup>74</sup> |
| *SORL1* | chr11:121604205:C:T | EFIGA | Missense | Endosomal degradation and recycling in neurons<sup>46</sup> |
| *SPPL2A* | chr15:50722170:A:T | AD-FBS | Missense | Involved in intramembrane cleavage of TMEM106B<sup>75</sup> |
| *TNIP1* | chr5:151052238:T:C | AD-FBS | Missense | Autoimmunity and tissue homeostasis via regulation of NFκB activation<sup>76</sup> |
| *TPCN1* | chr12:113293011:C:T<br>chr12:113288795:G:A | EFIGA<br>AD-FBS | Missense<br>Missense | Lysosomal TPCN inhibition ameliorates Aβ pathology<sup>77</sup> |
| *TREML2* | chr6:41198273:C:A | EFIGA | Missense | Hyperphosphorylation and aggregation of tau and induces activation of microglia<sup>78</sup> |
| *TSPOAP1* | chr17:58307871:C:G<br>chr17:58325608:C:T<br>chr17:58327896:G:A | EFIGA<br>EFIGA<br>EFIGA | Missense | Phosphatidylglycerol-regulated functions<sup>79</sup> |
| *UNC5CL* | chr6:41033992:C:T | EFIGA | Missense | Differential methylation related to Aβ plaques<sup>80</sup> |
| *USP6NL* | chr10:11463631:G:A | AD-FBS | Missense | Acts as a GTPase-activating protein for Ras-related protein<sup>81</sup> |
| *WDR81* | chr17:1732823:G:A<br>chr17:1728303:G:C | EFIGA<br>AD-FBS | Missense<br>Missense | Promotes the autophagic clearance of protein aggregates<sup>82</sup> |

AD-FBS: Alzheimer's Disease - National Institute on Aging Family Based Study, EFIGA: Estudio Familiar de la Genética de la Enfermedad.

### 3.2 *APOE* co-segregation

*APOE-ε4* co-segregated with LoF variants in *ABCA1, DOC2A*, and SLC26A1/IDUA in four of the white, non-Hispanic families, but there was no co-segregation in Hispanic families with any LoF variant. Missense variants in nine genes co-segregated with *APOE-ε4* including: *ABCA1, AKAP9, ANK3, CTSB, EED, PRDM7, SNX1, USP6NL, WDR81* in the AD-FBS families, while in the EFIGA families *APOE-ε4* segregated with missense variants in *ADAMTS20, AKAP9, ANK3, CR1, SORL1* and *TSPOAP1.* The presence of *APOE ε4* did not affect age at onset of dementia (Table 3). *APOE-e4* was the only variant segregating 93 (22.6%) of families, Among the AD-FBS cohort, 64 (32.5%) families had only segregation with *APOE-ε4*, and in the EFIGA cohort, 29 (13.5%) families had only *APOE-ε4* segregation.

**Table 3:**
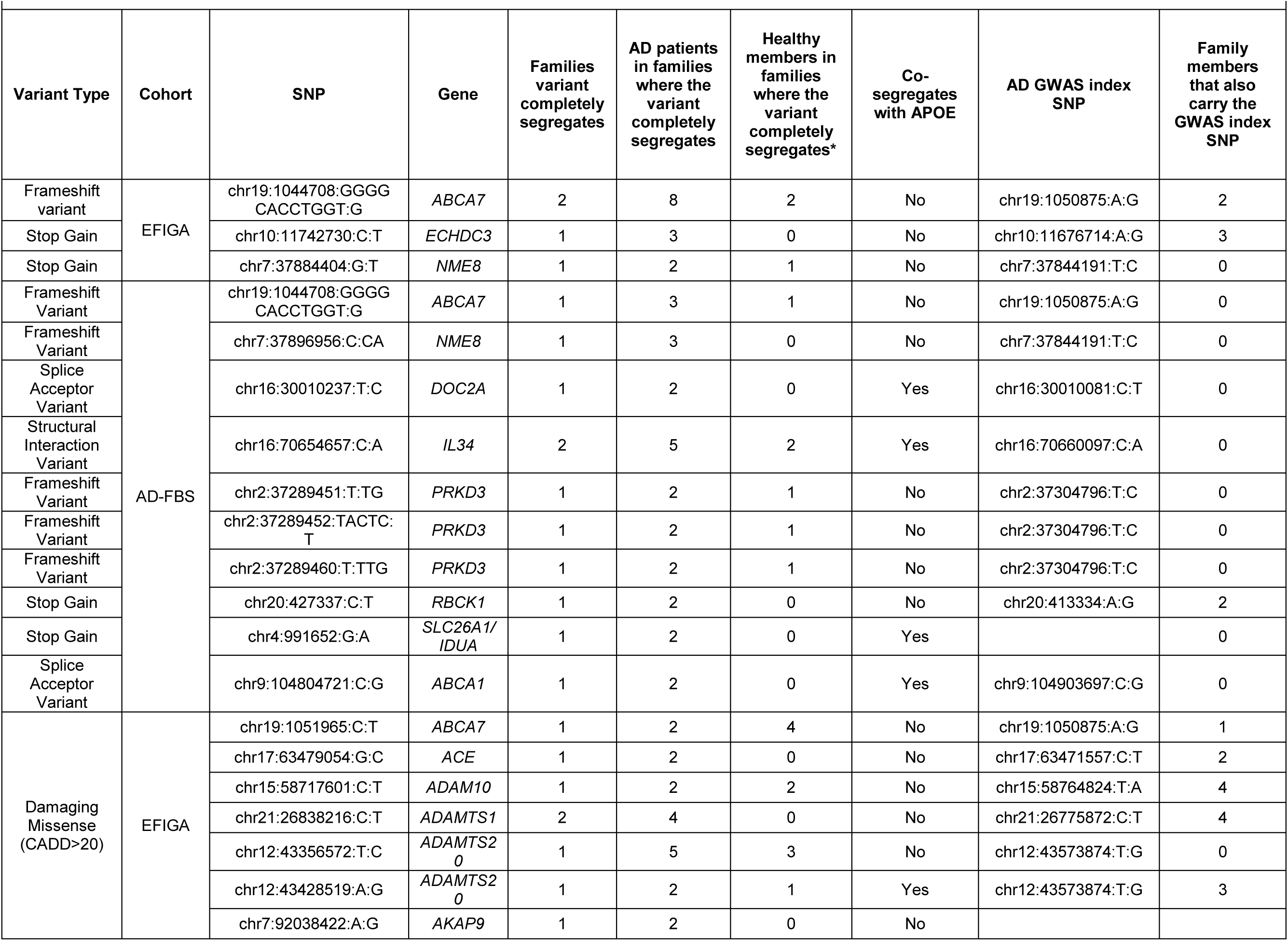
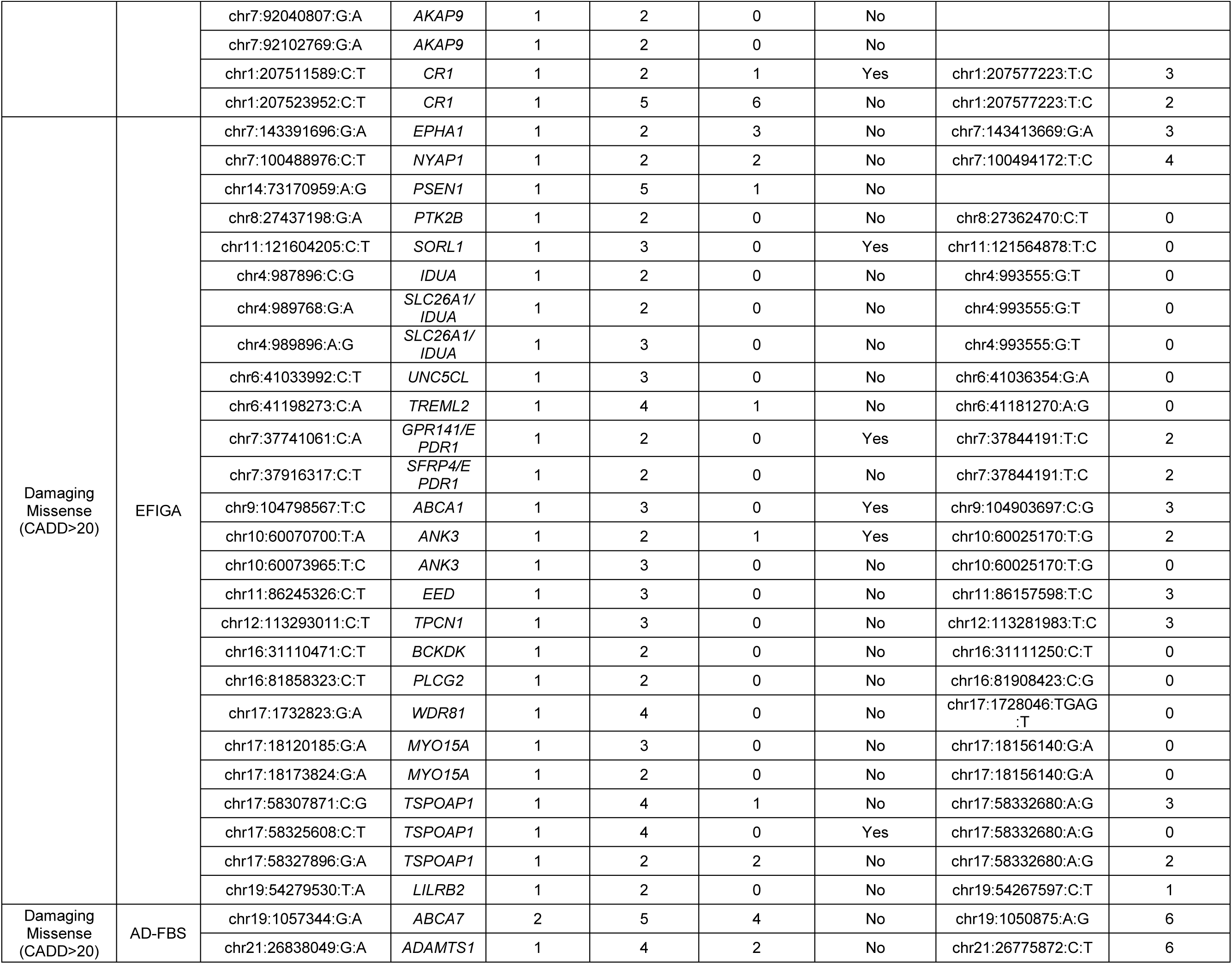
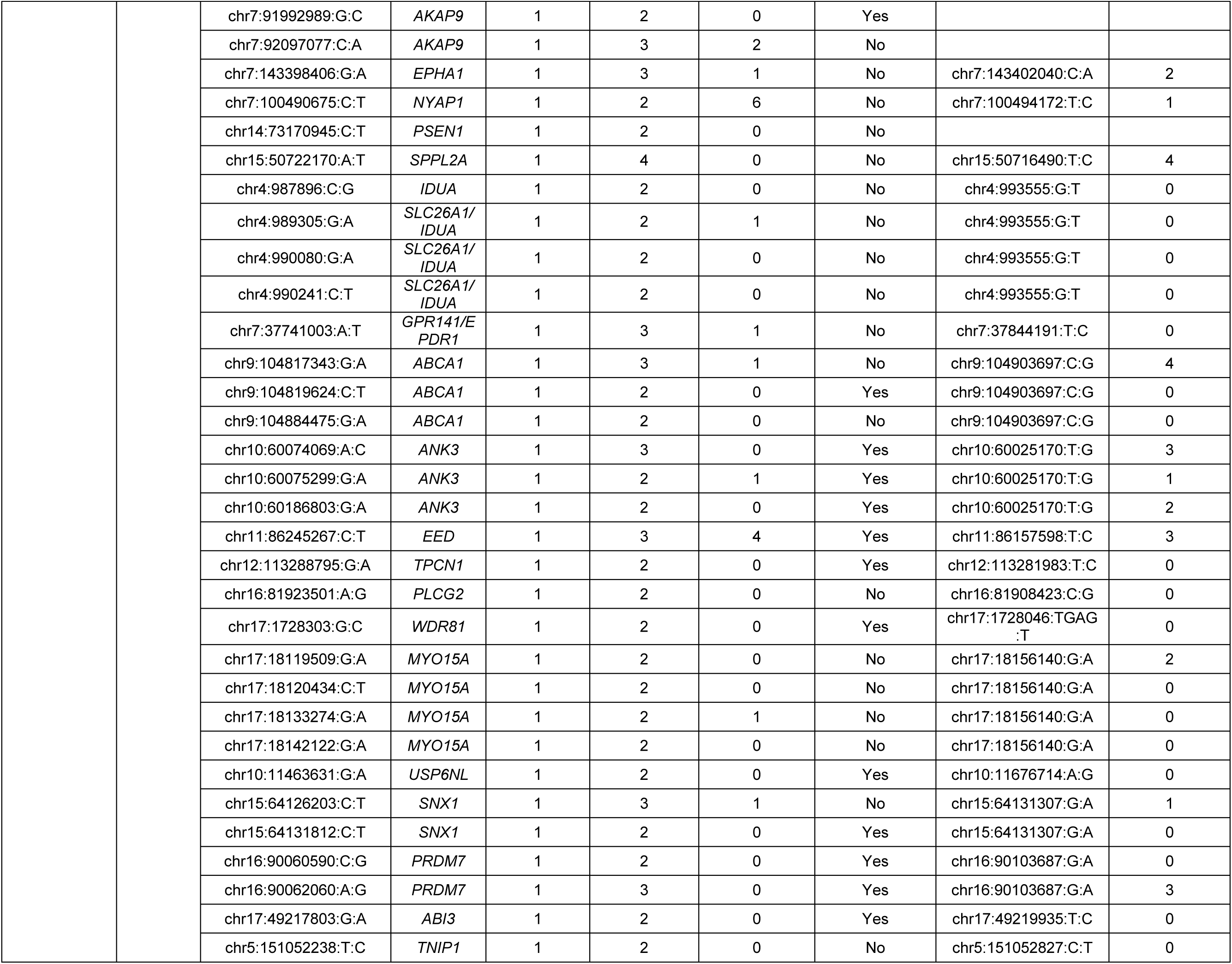
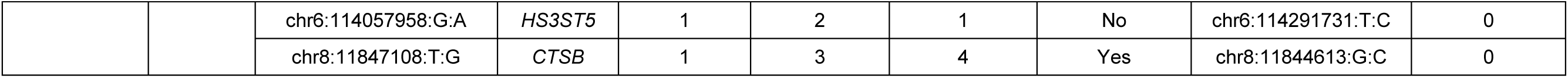
Details of variants that are completely segregating in at least one family.

### 3.3 Linkage disequilibrium (LD) between common and rare variants

Thirty-three (43.4%) of the 76 damaging missense variants or LoF variants were in linkage disequilibrium with the most significant common variant from the GWAS^8^ or with any of the four other candidate genes. Because LD is sensitive to allele frequency, we anticipated that the likelihood of a common and rare variant being in LD would be very low. Thus, we also investigated whether the most significant common variant from the GWAS data co-segregated with any rare variant we identified. Within AD-FBS families, we identified one LoF variant in *RBCK1* and ten damaging missense variants that co-segregated with the common variant (Table 3). Within EFIGA families, 15 damaging missense variants and one LoF variant completely co-segregated with the common variants (Table 3).

In 64 (32.5%) of the AD-FBS families and 29 (13.6%) of the EFIGA families, there were no segregating variants in any of the GWAS loci and *APOE-ε4* was the only risk allele segregating in these families. Among the AD-FBS cohort, 99 (50.3%) families had no rare variants identified and were not found to have *APOE-ε4* accounting for the segregation within the families. Similarly, 149(69.6%) of the EFIGA families had no rare variants identified and did not have *APOE-ε4* segregation.

## 4.0 DISCUSSION

Using existing data from whole genome sequencing in two groups of families multiply affected by late onset fAD we identified rare, potentially pathogenic, segregating variants embedded in or near risk loci reported from a large GWAS of unrelated individuals^8^, present in Mendelian forms of early onset fAD,^2–5^or identified by small sequencing investigations (e.g. *AKAP9).*^21^ We found numerous coding variants prior to restricting the analyses to damaging missense variants and LoF variants at these loci. In 38 AD-FBS families, variants in 29 genes were identified as fully segregating in all affected family members and younger relatives at risk (Table 3). Similarly, in 34 EFIGA families, variants in 31 genes were observed segregating. The highest frequency of segregating variants was found in *ABCA1, AKAP9, ANK3, MYO15A, and SLC26A1* in both groups of families. We also found no segregating variants within the GWAS loci and no evidence of *APOE-ε4* segregation in 60.3% of the families. This clearly indicates that an unbiased search for additional pathogenic variants, including structural variants should be undertaken to fully explain the genetic basis of fAD.

Genome wide array studies were designed using common genetic variation across the genome to identify loci that are associated with a trait or disease. Most are common variants that confer small effects regarding the proportion of the trait or disease risk. Rare variants are defined by allele frequency of less than 0.01. This is an arbitrary criterion, but it is well-accepted in the field and based on the observed effect sizes for variants at different MAF thresholds.^22^ Rare variants generally have a larger effect size compared with common variants, and they are more suitable for functional genomic and mechanistic analyses investigating them as potential drug targets. They can also be used for genetic counseling or as putative biomarkers to support a personalized medicine.^23^

Determining whether the rare variants identified in these two cohorts were in LD with the common variants from the GWAS loci can be difficult because the much higher frequency of the significant common variant compared with the rare variant. As an alternative we determined the frequency by which the significant common variant in the GWAS co-segregated with the rare variant. Among the AD-FBS families one (10%) LoF variants and 10 (41.7%) missense variants co-segregated with the significant common variant, compared with 2(66.7%) LoF variants and 16 (57.1%) missense variants co-segregated with the significant common variant in EFIGA families (Table 2).

Rare variants were found in *ABCA1, AKAP9,* and *ABCA7* which are well established as genes involved in AD. A variant in *ABCA1*, an ATP-binding cassette transporter A1, was identified in a large exome sequencing study^24^ of AD, and associated with reduced ApoE levels and decreased clearance of Aβ.^25^ This specific variant was not observed in the family data but several rare variants in *ABCA1* co-segregated with chr9: 104794495 (Table 3). Rare variants in A-kinase anchoring protein 9, *AKAP9*, were described in a small cohort of black, non-Hispanic individuals.^21^ Subsequently, a rare variant (p.Arg434Trp) was observed in two large families from the Dominican Republic.^13^ *AKAP9* contains 51 exons and is a binding protein that connects protein kinase to intracellular locations promoting cAMP related activation of PKA, which, in turn, enhances tau phosphorylation.^26^ This process is one of the major findings in postmortem AD brains. Although only *ABCA7* was first identified using common variants in a GWAS.^27^ The ATP-binding cassette, sub-family A, member 7 (*ABCA7*) gene, has both common risk variants with functional consequences and rare, damaging coding variants. *ABCA7* is a large gene with 47 exons encoding a transmembrane transporter protein. *ABCA7* is expressed in brain, especially in microglia, neurons, endothelial cells, pericytes where it may help degrade Aβ.^28^ It may also have an important role in lipid metabolism. Common variants in *ABCA7* associated with AD have been observed in black and white, non-Hispanic individuals as well as in Hispanics.^27; 28^ There have also been family data where premature termination codons have been found in *ABCA7.* ^29^ *ANK3* gene encodes for multiple isoforms of the scaffold protein, Ankyrin G, which is involved in protein interactions and neuronal functions. Mutations in ANK3 alter neuronal excitability and connectivity by altering ion channels and membrane proteins.^30^ *ANK1* has found to be differentially methylated in AD. Several variants in *MYO15A*, which encodes a myosin involved in actin organization in mechanosensory hair bundles.^31^ Actin is involved in retromer function which occurs in several neurodegenerative diseases including AD. SLC26A1 encodes a protein which acts as a transporter for ions and organic substrates ^32^ Variants in SLC26A1 may disrupt the function of the gene leading to oxidative stress, inflammation and metabolic dysfunction.^33^

Rare variants provide hypothesis free evidence for causality and can be targets for functional analyses to understand disease mechanisms.^23^ Thus, while variants in several other AD genetic loci were less frequent, they are no less important. Several lines of evidence have implicated the angiotensin converting enzyme, *ACE* in AD.^34; 35^ *ACE* encodes ACE-1, a rate-limiting enzyme in the cRAS pathway which generates disease-associated angiotensin II (Ang II) but also degrades and facilitates the clearance of Aβ. The gene encoding disintegrin-like and metalloproteinase with thrombospondin type 1 (*ADAMTS1*) is a member of the ADAMTS protein family. This family of genes are distinguished from *ADAM10* based on the number of copies of the thrombospondin 1-like repeats. Both are involved in extracellular matrix damage and repair.^36^ *ADAMTS1* is thought to be involved in hydrolysis of APP and consequently decreases Aβ generation through inhibiting β-secretase-mediated cleavage. Clinically the gene may also affect cognitive ability by its effect on hippocampal amyloid processing. *CR1*, which encodes a complement receptor, regulating the complement cascade to clear cellular debris, including Aβ.^37^ *ECHDC3* encodes an enzyme, enoyl CoA hydratase domain containing, and is a gene known to be involved in plasma lipids and other lipid related traits. Its expression is altered in the brains of patients with AD.^38^ *EPHA1* belongs to the ephrin receptor subfamily of the protein-kinase family, and these receptors have been involved in development of the nervous system. In an early GWAS of AD, the Alzheimer’s Disease Genetics Consortium identified a common variant in *EPHA1* associated with ADGene Ontology annotations related to this gene include transferase activity, transferring phosphorus-containing groups and protein tyrosine kinase activity.^39^ Variants in Neuronal Tyrosine Phosphorylated Phosphoinositide-3-Kinase Adaptor 1, *NYAP1*, appear to regulate immunoglobulin-like receptors beta and alpha (PILRB and PILRA), ^40^ but it is unclear how these variants contribute to AD pathogenesis. Presenilin 1, *PSEN1,* a gene associated with early onset AD, has been previously observed in late onset, fAD^41; 42^ and it encodes the catalytic sub-unit of α-secretase involved in Aβ processing.

Variants in Protein Tyrosine Kinase 2 Beta, *PTK2B*, are thought to disrupt the normal suppression of tau phosphorylation and the effects of tauopathy thereby adding to the pathogenesis of AD.^43–45^ Sortilin-related receptor with A-type repeats (SORLA or SORL1) is a sorting receptor in which variants impair processing of APP to soluble (s)APP and to the amyloid beta-peptide. SORL1 is also involved in endosomal degradation of Aβ and recycling in neurons.^24; 46^ Finally, a common variant in signal peptide peptidase-like 2A*, SPPL2A,* was identified in a large genome wide array, and the gene is thought to be part of the immune response.^8; 47^ We annotated the function of other genes with segregating variants in Table 2.

Rare variants have distinctive features, low likelihood of linkage disequilibrium with other variants in the region, more likely to affect gene function or expression, and a more direct role in diseases.^23^ Efforts to discover associations driven by low-frequency, rare variants through genome sequencing efforts provide refinements to estimates of disease heritability.^48^ Identifying rare variants in protein-coding regions through genome sequencing would also greatly facilitate annotating genes associated with complex disease and describing the functional consequences of human sequence variation.

The reduction in costs for genetic testing will also make it more feasible to include in genome or exome sequencing in clinical practice. Strategies to identify damaging variants will soon be in place and clinicians and counselors will need to be able to identify and explain these results to patients and families. Families generally want to know the diagnostic relevance of our findings and the potential recurrence risk to other family. Therefore, a compelling argument can be made to accelerate efforts to identify rare variants in fAD because of the relative importance of turning these discoveries into meaningful clinical and biological progress.

## Data Availability

All clinical data are available upon request by specific projects included in this manuscript. All genetic data are available from the NIA Genetics of Alzheimer's Disease (NIAGADS) Data Sharing Service (DSS) hosts and shares data generated from the ADSP with the research community.

https://www.neurology.columbia.edu/research/research-centers-and-programs/alzheimers-disease-research-center-adrc/investigators/investigator-resources.

https://dss.niagads.org/documentation/data-application-and-submission/application-instructions

## Acknowledgements

The authors would like to thank the project managers at our respective sites for their efforts in recruitment and retention of these families.

## Consent Statement

**NIA AD-FBS.** Every institution was required to have their local Investigational Review Board and approve the consent form for this investigation. Thus, all individuals provided informed consent.

**EFIGA.** The Dominican Republic has a centralized Investigational Review Board. They approved the consent form used in this investigation for families living in the Dominican Republic. Thus, all individuals provided informed consent.

