## Supplemental Tables 1, 2 and 3 for "Missense and Loss of Function Variants at GWAS Loci in Familial Alzheimer’s Disease"

**Supplementary Table 1. Genes included in the segregation analysis**

| Chromosome | Gene | Number of variants screened for the segregation |
|---|---|---|
| 9 | *ABCA1* | 88 |
| 19 | *ABCA7* | 201 |
| 17 | *ABI3* | 17 |
| 17 | *ACE/CTD-2501B8.1* | 62 |
| 15 | *ADAM10* | 14 |
| 2 | *ADAM17* | 100 |
| 21 | *ADAMTS1* | 48 |
| 12 | *ADAMTS20* | 95 |
| 7 | *AKAP9* | 158 |
| 10 | *ANK3* | 145 |
| 5 | *ANKH* | 13 |
| 15 | *APH1B* | 26 |
| 19 | *APOE* | 22 |
| 21 | *APP* | 21 |
| 16 | *BCKDK* | 25 |
| 2 | *BIN1* | 16 |
| 10 | *BLNK* | 29 |
| 20 | *CASS4* | 47 |
| 6 | *CD2AP* | 32 |
| 4 | *CLNK* | 34 |
| 8 | *CLU* | 24 |
| 1 | *CR1* | 105 |
| 8 | *CTSB* | 42 |
| 15 | *CTSH* | 21 |
| 16 | *DOC2A* | 31 |
| 11 | *EED/PICALM* | 37 |
| 7 | *EPDR1/NME8/GPR141* | 124 |
| 7 | *EPHA1* | 70 |
| 14 | *FERMT2* | 21 |
| 16 | *FOXF1* | 14 |
| 5 | *FST* | 9 |
| 6 | *HLA-DQA1/HLA-DRB1* | 138 |
| 6 | *HS3ST5* | 18 |
| 7 | *ICA1* | 20 |
| 4 | *IDUA/SLC26A1* | 850 |
| 16 | *IL34* | 46 |
| 2 | *INPP5D* | 46 |
| 16 | *IQCK* | 15 |
| 7 | *JAZF1* | 4 |
| 19 | *KLF16* | 18 |
| 19 | *LILRB2* | 156 |
| 16 | *MAF* | 40 |
| 5 | *MEF2C* | 9 |

| | | |
|---|---|---|
| 3 | *MME* | 47 |
| 11 | *MS4A2* | 19 |
| 11 | *MS4A4A* | 25 |
| 17 | *MYO15A* | 219 |
| 2 | *NCK2* | 16 |
| 8 | *NDUFAF6* | 22 |
| 16 | *PLCG2* | 76 |
| 10 | *PLEKHA1* | 24 |
| 16 | *PRDM7* | 70 |
| 2 | *PRKD3* | 43 |
| 14 | *PSEN1* | 11 |
| 1 | *PSEN2* | 31 |
| 8 | *PTK2B* | 42 |
| 5 | *RASGEF1C* | 15 |
| 20 | *RBCK1* | 24 |
| 17 | *SCIMP* | 12 |
| 8 | *SHARPIN* | 30 |
| 19 | *SIGLEC11* | 127 |
| 14 | *SLC24A4* | 38 |
| 20 | *SLC2A4RG* | 28 |
| 15 | *SNX1* | 30 |
| 11 | *SORL1* | 100 |
| 1 | *SORT1* | 35 |
| 7 | *SPDYE3* | 38 |
| 11 | *SPI1* | 9 |
| 15 | *SPPL2A* | 23 |
| 7 | *TMEM106B* | 11 |
| 5 | *TNIP1* | 37 |
| 12 | *TPCN1* | 33 |
| 6 | *TREM2/ORAD1* | 19 |
| 6 | *TREML2* | 37 |
| 17 | *TSPOAP1* | 118 |
| 7 | *UMAD1* | 15 |
| 6 | *UNC5CL* | 34 |
| 10 | *USP6NL/ECHDC3* | 67 |
| 2 | *WDR12/CARF* | 38 |
| 17 | *WDR81* | 129 |
| 17 | *WNT3* | 6 |

**Supplementary Table 2. Allele frequencies of segregating variants in the ADSP cohort**

| Variant Type |
|---|
| Frameshift variant |
| Frameshift Variant |
| Splice Acceptor Variant |
| Damaging Missense (CADD>20)<br>Damaging Missense (CADD>20)<br>Damaging Missense (CADD>20 |
| Damaging Missense (CADD>20) |
| Damaging Missense (CADD>20) |

| Cohort | SNP | Gene | Minor allele frequency in ADSP cohort | | |
|---|---|---|---|---|---|
| EFIGA | chr19:1044708:GGGG CACCTGGT:G | *ABCA7* | 0.00125 | 0.00103 | 0.00098 |
| AD-FBS | chr19:1044708:GGGG CACCTGGT:G | *ABCA7* | 0.00125 | 0.00103 | 0.00098 |
| EFIGA | chr16:30010237:T:C | *DOC2A* | 0.00069 | 0.00081 | 0.00066 |
| | chr15:58717601:C:T | *ADAM10* | 0.00074 | 0.00109 | 0.0009 |
| | chr12:43356572:T:C | *ADAMTS20* | 0.00162 | 0.00121 | 0.00115 |
| | chr12:43428519:A:G | *ADAMTS20* | 0.00157 | 0.00174 | 0.00152 |
| | chr7:92040807:G:A | *AKAP9* | 0.00028 | 0.00075 | 0.00055 |
| | chr1:207511589:C:T | *CR1* | 0.00028 | 0.00019 | 0.00018 |
| | chr1:207523952:C:T | *CR1* | 0.00028 | 0.00031 | 0.00029 |
| | chr7:143391696:G:A | *EPHA1* | 0.00005 | 0.00044 | 0.00029 |
| EFIGA | chr8:27437198:G:A | *PTK2B* | 0.00009 | 0.00019 | 0.00017 |
| | chr11:121604205:C:T | *SORL1* | 0.00023 | 0.00047 | 0.00036 |
| | chr6:41033992:C:T | *UNC5CL* | 0.00055 | 0.00072 | 0.00051 |
| | chr6:41198273:C:A | *TREML2* | 0.00069 | 0.00096 | 0.0008 |
| EFIGA | chr10:60073965:T:C | *ANK3* | 0.00037 | 0.00037 | 0.00035 |
| | chr17:1732823:G:A | *WDR81* | 0.00042 | 0.00044 | 0.00043 |
| | chr17:58307871:C:G | *TSPOAP1* | 0.00125 | 0.00165 | 0.00123 |
| | chr17:58325608:C:T | *TSPOAP1* | 0.00102 | 0.00087 | 0.00082 |
| | chr19:54279530:T:A | *LILRB2* | 0.00166 | 0.00143 | 0.00141 |
| AD-FBS | chr19:1057344:G:A | *ABCA7* | 0.003 | 0.00389 | 0.00375 |
| | chr9:104817343:G:A | *ABCA1* | 0.00005 | 0.00006 | 0.00004 |
| | chr17:18119509:G:A | *MYO15A* | 0.00074 | 0.00078 | 0.00064 |
| AD-FBS | chr16:90060590:C:G | *PRDM7* | 0.00023 | 0.00093 | 0.00076 |
| | chr16:90062060:A:G | *PRDM7* | 0.00005 | 0.00006 | 0.00004 |
| | chr5:151052238:T:C | *TNIP1* | 0.00194 | 0.00227 | 0.00203 |
| | chr8:11847108:T:G | *CTSB* | 0.0032 | 0.00393 | 0.00376 |

1

1
1
1
1
1
1
1
1
1
1
1
1
1
1
1
1
1
1
1
1
1
1
1
1
1

#### Table 3. Variants- Incomplete Segregation in Both Datasets

| SNP | EFFECT | GENE | Total Carriers subjects |
|---|---|---|---|
| chr19:1041509:G:A | splice_acceptor_variant&intron_variant | ABCA7 | 6 |
| chr7:92022822:CA:C | frameshift_variant | AKAP9 | 5 |
| chr11:60090358:TG:T | frameshift_variant | MS4A2 | 7 |
| chr2:202983584:C:CA | frameshift_variant | CARF | 15 |
| chr20:417604:G:A | structural_interaction_variant | RBCK1 | 8 |
| chr19:54279447:C:CG | frameshift_variant | LILRB2 | 8 |
| chr17:18119497:GGC:G | frameshift_variant | MYO15A | 3 |
| chr17:5223369:TGAGGCCCA:T | frameshift_variant | SCIMP | 4 |
| chr2:105881962:C:T | structural_interaction_variant | NCK2 | 1 |
| chr4:10528096:T:G | splice_acceptor_variant&intron_variant | CLNK | 1 |
| chr4:10558454:T:C | splice_acceptor_variant&intron_variant | CLNK | 1 |
| chr4:1003541:T:TCCCCC | splice_acceptor_variant&intron_variant | IDUA | 1 |
| chr7:37741212:AG:A | frameshift_variant | GPR141 | 1 |
| chr7:37864394:T:TA | frameshift_variant | NME8 | 1 |
| chr7:37864412:TAAAAG:T | frameshift_variant&splice_donor_variant& | NME8 | 1 |
| chr7:37867795:GA:G | frameshift_variant | NME8 | 3 |
| chr7:37897068:CTT:C | frameshift_variant | NME8 | 1 |
| chr7:100309094:G:A | stop_gained | SPDYE3 | 3 |
| chr7:100311812:TTGGA:T | frameshift_variant | SPDYE3 | 1 |

#### AD-FBS LoF variants- Incomplete Segregation

| SNP | EFFECT | GENE | Total Carriers subjects |
|---|---|---|---|
| chr10:11763413:C:T | stop_gained | ECHDC3 | 7 |
| chr11:60094000:G:T | stop_gained | MS4A2 | 5 |
| chr16:90075897:C:CT | frameshift_variant | PRDM7 | 1 |
| chr17:18138933:C:G | stop_gained | MYO15A | 1 |
| chr4:10566062:C:T | stop_gained | CLNK | 1 |
| chr4:1002747:G:A | structural_interaction_variant | IDUA | 12 |
| chr4:1003563:C:T | structural_interaction_variant | IDUA | 1 |
| chr4:1004011:G:C | splice_acceptor_variant&intron_variant | IDUA | 1 |
| chr4:989203:TC:T | frameshift_variant | SLC26A1 | 4 |

| | | | |
|---|---|---|---|
| chr7:37740394:A:AT | frameshift_variant&start_lost | GPR141 | 1 |
| chr7:37740396:G:C | start_lost | GPR141 | 1 |

### EFIGA CADD > 20 variants - Incomplete segregation

| SNP | EFFECT | GENE | Total Carriers subjects |
|---|---|---|---|
| chr19:1041972:T:G | missense_variant&splice_region_variant | ABCA7 | 7 |
| chr19:1043794:C:T | missense_variant | ABCA7 | 5 |
| chr19:1056089:G:T | missense_variant | ABCA7 | 4 |
| chr17:63477303:C:T | missense_variant | ACE | 5 |
| chr17:63481683:C:T | missense_variant | ACE | 4 |
| chr17:63481708:C:T | missense_variant | ACE | 8 |
| chr17:63494394:C:A | missense_variant | TD-2501B8 | 11 |
| chr21:26837670:T:C | missense_variant | ADAMTS1 | 9 |
| chr7:92001425:A:G | missense_variant | AKAP9 | 9 |
| chr7:92001542:A:T | missense_variant | AKAP9 | 6 |
| chr7:92070085:T:G | missense_variant | AKAP9 | 11 |
| chr7:92083494:G:A | missense_variant | AKAP9 | 11 |
| chr7:92097077:C:A | missense_variant | AKAP9 | 4 |
| chr21:26000030:C:T | missense_variant | APP | 8 |
| chr21:26021879:T:A | missense_variant | APP | 4 |
| chr20:56451910:A:G | missense_variant | CASS4 | 19 |
| chr20:56458449:G:C | missense_variant | CASS4 | 7 |
| chr8:27606389:C:T | missense_variant | CLU | 5 |
| chr1:207580249:C:T | missense_variant | CR1 | 6 |
| chr7:143399326:G:A | missense_variant | EPHA1 | 4 |
| chr2:233170072:G:T | missense_variant | INPP5D | 5 |
| chr16:19763870:C:T | missense_variant | IQCK | 4 |
| chr8:95052195:G:A | missense_variant | NDUFAF6 | 4 |
| chr7:37894471:A:C | missense_variant | NME8 | 12 |
| chr14:92443472:G:A | missense_variant&splice_region_variant | SLC24A4 | 14 |
| chr2:9518204:A:C | missense_variant | ADAM17 | 1 |
| chr2:9527894:C:A | missense_variant | ADAM17 | 5 |
| chr2:37253209:G:C | missense_variant | PRKD3 | 1 |
| chr2:37256677:C:T | missense_variant | PRKD3 | 1 |
| chr2:37279788:T:C | missense_variant | PRKD3 | 1 |
| chr2:37316249:T:C | missense_variant | PRKD3 | 1 |
| chr2:37316424:G:A | missense_variant | PRKD3 | 6 |
| chr2:202952598:C:T | missense_variant | CARF | 2 |
| chr3:155115014:A:G | missense_variant | MME | 1 |
| chr3:155142009:T:A | missense_variant | MME | 2 |

| | | | |
|---|---|---|---|
| chr3:155168594:A:G | missense_variant | MME | 3 |
| chr3:155172203:C:A | missense_variant | MME | 2 |
| chr4:987174:C:G | missense_variant | IDUA | 1 |
| chr4:988981:A:G | missense_variant | SLC26A1 | 2 |
| chr4:989033:C:A | missense_variant | SLC26A1 | 11 |
| chr4:989092:C:G | missense_variant | SLC26A1 | 7 |
| chr4:989428:C:T | missense_variant | SLC26A1 | 16 |
| chr4:989861:C:T | missense_variant | SLC26A1 | 1 |
| chr4:989965:G:A | missense_variant | SLC26A1 | 6 |
| chr4:991538:C:T | missense_variant | SLC26A1 | 6 |
| chr4:991648:C:G | missense_variant | SLC26A1 | 13 |
| chr4:1003582:G:C | missense_variant | IDUA | 2 |
| chr4:10490492:A:T | missense_variant | CLNK | 3 |
| chr4:10501312:G:A | missense_variant | CLNK | 4 |
| chr4:10598015:C:T | missense_variant | CLNK | 5 |
| chr5:14711223:C:T | missense_variant | ANKH | 8 |
| chr5:14751230:C:T | missense_variant | ANKH | 1 |
| chr5:151036890:G:A | missense_variant | TNIP1 | 11 |
| chr5:151042607:C:T | missense_variant | TNIP1 | 2 |
| chr5:151045935:C:G | missense_variant | TNIP1 | 2 |
| chr5:151052247:G:T | missense_variant | TNIP1 | 1 |
| chr6:41033103:G:A | missense_variant | UNC5CL | 10 |
| chr6:114057325:G:A | missense_variant | HS3ST5 | 1 |
| chr6:114058083:C:T | missense_variant | HS3ST5 | 1 |
| chr7:8221365:A:C | missense_variant | ICA1 | 2 |
| chr7:27832847:A:G | missense_variant | JAZF1 | 1 |
| chr7:37907573:G:T | missense_variant | SFRP4 | 2 |
| chr7:37948875:T:C | missense_variant | EPDR1 | 4 |
| chr7:37950287:C:T | missense_variant | EPDR1 | 19 |
| chr7:37950326:C:T | missense_variant | EPDR1 | 4 |
| chr7:37950358:G:A | missense_variant | EPDR1 | 5 |
| chr7:100319970:G:C | missense_variant | SPDYE3 | 1 |
| chr7:100319974:G:C | missense_variant | SPDYE3 | 3 |
| chr8:11849163:G:A | missense_variant&splice_region_variant | CTSB | 1 |
| chr8:144098934:T:C | missense_variant | SHARPIN | 4 |
| chr9:104788006:T:C | missense_variant | ABCA1 | 1 |
| chr9:104788041:G:A | missense_variant | ABCA1 | 3 |
| chr9:104791982:C:T | missense_variant | ABCA1 | 10 |
| chr9:104792796:T:G | missense_variant | ABCA1 | 1 |
| chr9:104816297:C:T | missense_variant | ABCA1 | 3 |
| chr9:104816337:C:T | missense_variant | ABCA1 | 3 |
| chr9:104889159:T:C | missense_variant | ABCA1 | 2 |
| chr10:11463250:C:A | missense_variant | USP6NL | 1 |
| chr10:60042719:C:T | missense_variant | ANK3 | 14 |
| chr10:60063239:G:A | missense_variant | ANK3 | 4 |
| chr10:60070193:T:C | missense_variant | ANK3 | 13 |
| chr10:60073926:C:T | missense_variant | ANK3 | 2 |

| | | | |
|---|---|---|---|
| chr10:60075875:G:A | missense_variant | ANK3 | 3 |
| chr10:60076308:A:G | missense_variant | ANK3 | 1 |
| chr10:60082736:A:G | missense_variant&splice_region_variant | ANK3 | 2 |
| chr10:60172376:G:A | missense_variant | ANK3 | 13 |
| chr10:60181352:C:A | missense_variant | ANK3 | 1 |
| chr10:60389436:G:A | missense_variant | ANK3 | 2 |
| chr10:122412965:C:T | missense_variant | PLEKHA1 | 3 |
| chr10:122415920:G:A | missense_variant | PLEKHA1 | 3 |
| chr10:122429900:G:A | missense_variant | PLEKHA1 | 1 |
| chr10:122429922:C:T | missense_variant | PLEKHA1 | 1 |
| chr15:64096021:C:T | missense_variant | SNX1 | 3 |
| chr15:64126084:C:G | missense_variant | SNX1 | 3 |
| chr15:64131765:G:A | missense_variant | SNX1 | 2 |
| chr15:64131787:G:T | missense_variant | SNX1 | 2 |
| chr15:64136334:A:C | missense_variant | SNX1 | 1 |
| chr15:78929473:A:G | missense_variant | CTSH | 1 |
| chr15:78931505:G:C | missense_variant&splice_region_variant | CTSH | 1 |
| chr15:78932389:C:T | missense_variant | CTSH | 1 |
| chr16:30006869:C:T | missense_variant | DOC2A | 4 |
| chr16:70660045:C:G | missense_variant | IL34 | 6 |
| chr16:70660087:C:T | missense_variant | IL34 | 1 |
| chr16:81907750:G:C | missense_variant | PLCG2 | 2 |
| chr16:81923501:A:G | missense_variant | PLCG2 | 1 |
| chr16:81931581:C:T | missense_variant | PLCG2 | 2 |
| chr17:1727354:C:T | missense_variant | WDR81 | 3 |
| chr17:1730401:G:A | missense_variant | WDR81 | 1 |
| chr17:1730833:C:T | missense_variant | WDR81 | 1 |
| chr17:1733760:C:T | missense_variant | WDR81 | 2 |
| chr17:1733832:G:A | missense_variant | WDR81 | 4 |
| chr17:1733881:G:A | missense_variant | WDR81 | 1 |
| chr17:1735623:C:T | missense_variant | WDR81 | 5 |
| chr17:18118886:G:A | missense_variant | MYO15A | 7 |
| chr17:18119069:T:C | missense_variant | MYO15A | 2 |
| chr17:18119509:G:A | missense_variant | MYO15A | 12 |
| chr17:18119677:G:A | missense_variant | MYO15A | 2 |
| chr17:18120430:C:T | missense_variant | MYO15A | 3 |
| chr17:18120434:C:T | missense_variant | MYO15A | 1 |
| chr17:18121256:C:G | missense_variant | MYO15A | 1 |
| chr17:18133328:T:C | missense_variant | MYO15A | 2 |
| chr17:18138863:C:T | missense_variant | MYO15A | 7 |
| chr17:18148100:G:A | missense_variant | MYO15A | 6 |
| chr17:18148133:C:T | missense_variant | MYO15A | 3 |
| chr17:18153858:T:C | missense_variant | MYO15A | 5 |
| chr17:18155414:G:A | missense_variant | MYO15A | 2 |
| chr17:18159308:G:C | missense_variant | MYO15A | 3 |
| chr17:18159677:A:C | missense_variant&splice_region_variant | MYO15A | 1 |
| chr17:18161408:C:T | missense_variant | MYO15A | 26 |

| | | | |
|---|---|---|---|
| chr17:18161436:T:C | missense_variant | MYO15A | 2 |
| chr17:18163800:C:T | missense_variant | MYO15A | 2 |
| chr17:18173823:C:T | missense_variant | MYO15A | 2 |
| chr17:49219932:C:T | missense_variant | ABI3 | 1 |
| chr17:49222558:C:A | missense_variant | ABI3 | 1 |
| chr17:58305442:C:T | missense_variant | TSPOAP1 | 2 |
| chr17:58307630:C:T | missense_variant | TSPOAP1 | 1 |
| chr17:58308579:G:A | missense_variant | TSPOAP1 | 5 |
| chr17:58308582:C:T | missense_variant | TSPOAP1 | 1 |
| chr17:58312155:C:T | missense_variant | TSPOAP1 | 1 |
| chr17:58312462:C:T | missense_variant | TSPOAP1 | 1 |
| chr17:58312671:C:T | missense_variant | TSPOAP1 | 1 |
| chr17:58322406:G:A | missense_variant | TSPOAP1 | 3 |
| chr17:58322982:G:A | missense_variant | TSPOAP1 | 5 |
| chr17:58324995:G:A | missense_variant | TSPOAP1 | 1 |
| chr17:58325653:G:A | missense_variant | TSPOAP1 | 2 |
| chr17:58325706:G:A | missense_variant | TSPOAP1 | 5 |
| chr19:49958880:C:T | missense_variant | SIGLEC11 | 2 |
| chr19:49960651:C:T | missense_variant | SIGLEC11 | 4 |
| chr19:54278817:A:G | missense_variant | LILRB2 | 1 |
| chr19:54278911:C:G | missense_variant | LILRB2 | 4 |
| chr20:417585:G:C | missense_variant | RBCK1 | 1 |

#### AD-FBS CADD > 20 variants - Incomplete segregation

| SNP | EFFECT | GENE | Total Carriers subjects |
|---|---|---|---|
| chr19:1055300:G:A | missense_variant | ABCA7 | 5 |
| chr17:63483485:C:G | missense_variant | ACE | 9 |
| chr7:91941109:G:C | missense_variant | AKAP9 | 4 |
| chr7:92097646:G:A | missense_variant | AKAP9 | 7 |
| chr7:92098165:A:T | missense_variant | AKAP9 | 11 |
| chr7:100488976:C:T | missense_variant | NYAP1 | 8 |
| chr14:92486772:C:T | missense_variant | SLC24A4 | 9 |
| chr11:121514222:A:C | missense_variant | SORL1 | 10 |
| chr11:121545284:T:A | missense_variant | SORL1 | 4 |
| chr11:121555188:G:A | missense_variant&splice_region_variant | SORL1 | 6 |
| chr10:11485832:C:A | missense_variant | USP6NL | 1 |
| chr10:60070484:A:T | missense_variant | ANK3 | 1 |
| chr10:60072134:A:T | missense_variant | ANK3 | 1 |
| chr10:60072489:C:T | missense_variant | ANK3 | 3 |
| chr10:60072558:C:G | missense_variant | ANK3 | 1 |

| | | | |
|---|---|---|---|
| chr10:60074900:G:A | missense_variant | ANK3 | 1 |
| chr10:60076142:G:A | missense_variant | ANK3 | 1 |
| chr10:60080569:T:C | missense_variant | ANK3 | 1 |
| chr10:60084679:T:C | missense_variant | ANK3 | 1 |
| chr10:60088199:T:C | missense_variant | ANK3 | 1 |
| chr10:60181340:C:T | missense_variant | ANK3 | 2 |
| chr10:60263863:T:C | missense_variant | ANK3 | 1 |
| chr10:96204068:A:G | missense_variant | BLNK | 2 |
| chr10:96227462:C:G | missense_variant | BLNK | 5 |
| chr10:122429922:C:T | missense_variant | PLEKHA1 | 1 |
| chr12:113293011:C:T | missense_variant | TPCN1 | 6 |
| chr15:63277669:C:T | missense_variant | APH1B | 1 |
| chr15:63305771:G:A | missense_variant | APH1B | 2 |
| chr15:64129919:C:A | missense_variant | SNX1 | 2 |
| chr15:64131870:T:A | missense_variant | SNX1 | 1 |
| chr15:64136878:C:A | missense_variant | SNX1 | 1 |
| chr16:30006245:G:A | missense_variant | DOC2A | 2 |
| chr16:31111999:A:T | missense_variant | BCKDK | 8 |
| chr16:70654584:G:T | missense_variant | IL34 | 1 |
| chr16:81883283:C:T | missense_variant | PLCG2 | 2 |
| chr16:81912698:C:G | missense_variant | PLCG2 | 3 |
| chr16:81923570:A:G | missense_variant | PLCG2 | 1 |
| chr16:90060451:C:T | missense_variant | PRDM7 | 3 |
| chr17:1726220:C:T | missense_variant | WDR81 | 1 |
| chr17:1726277:C:G | missense_variant | WDR81 | 1 |
| chr17:1726574:C:T | missense_variant | WDR81 | 2 |
| chr17:1727529:C:G | missense_variant | WDR81 | 1 |
| chr17:1728516:C:T | missense_variant | WDR81 | 1 |
| chr17:1733860:G:A | missense_variant | WDR81 | 1 |
| chr17:1735584:G:A | missense_variant | WDR81 | 4 |
| chr17:1737441:C:T | missense_variant | WDR81 | 2 |
| chr17:1737494:G:A | missense_variant | WDR81 | 1 |
| chr17:1737602:A:G | missense_variant | WDR81 | 1 |
| chr17:18119141:G:T | missense_variant | MYO15A | 2 |
| chr17:18120254:T:C | missense_variant | MYO15A | 1 |
| chr17:18136427:G:T | missense_variant | MYO15A | 3 |
| chr17:18136684:G:A | missense_variant&splice_region_variant | MYO15A | 4 |
| chr17:18138863:C:T | missense_variant | MYO15A | 3 |
| chr17:18142796:C:T | missense_variant | MYO15A | 1 |
| chr17:18148133:C:T | missense_variant | MYO15A | 6 |
| chr17:18150737:C:G | missense_variant | MYO15A | 1 |
| chr17:18151114:C:T | missense_variant | MYO15A | 1 |
| chr17:18156263:A:G | missense_variant | MYO15A | 1 |
| chr17:18172183:G:A | missense_variant | MYO15A | 3 |
| chr17:18173839:C:T | missense_variant | MYO15A | 4 |
| chr17:49217800:A:G | missense_variant | ABI3 | 1 |
| chr17:58308579:G:A | missense_variant | TSPOAP1 | 1 |

| | | | |
|---|---|---|---|
| chr17:58308770:T:C | missense_variant | TSPOAP1 | 1 |
| chr17:58309059:G:A | missense_variant | TSPOAP1 | 3 |
| chr17:58310152:C:T | missense_variant | TSPOAP1 | 1 |
| chr17:58312155:C:T | missense_variant | TSPOAP1 | 1 |
| chr17:58312462:C:T | missense_variant | TSPOAP1 | 1 |
| chr17:58316465:G:A | missense_variant | TSPOAP1 | 7 |
| chr17:58319270:G:C | missense_variant | TSPOAP1 | 1 |
| chr17:58322982:G:A | missense_variant | TSPOAP1 | 2 |
| chr19:49960389:C:T | missense_variant | SIGLEC11 | 1 |
| chr19:54278442:G:A | missense_variant | LILRB2 | 2 |
| chr1:109355392:G:C | missense_variant | SORT1 | 1 |
| chr20:428566:C:T | missense_variant | RBCK1 | 1 |
| chr2:9493755:T:C | missense_variant | ADAM17 | 1 |
| chr2:9497230:G:A | missense_variant | ADAM17 | 1 |
| chr2:9536773:C:A | missense_variant | ADAM17 | 1 |
| chr2:202884286:A:C | missense_variant | WDR12 | 1 |
| chr2:202952598:C:T | missense_variant | CARF | 9 |
| chr3:155140209:G:A | missense_variant | MME | 2 |
| chr3:155142073:A:G | missense_variant | MME | 1 |
| chr4:989033:C:A | missense_variant | SLC26A1 | 8 |
| chr4:989062:C:T | missense_variant | SLC26A1 | 1 |
| chr4:991522:C:T | missense_variant | SLC26A1 | 1 |
| chr4:1000668:A:C | missense_variant | IDUA | 2 |
| chr4:10490492:A:T | missense_variant | CLNK | 1 |
| chr4:10501311:C:T | missense_variant | CLNK | 1 |
| chr4:10542022:C:A | missense_variant&splice_region_variant | CLNK | 2 |
| chr5:14749204:G:A | missense_variant | ANKH | 2 |
| chr5:151039190:C:A | missense_variant | TNIP1 | 1 |
| chr5:180111505:C:T | missense_variant | RASGEF1C | 1 |
| chr6:41030401:C:T | missense_variant | UNC5CL | 4 |
| chr6:41032063:G:A | missense_variant | UNC5CL | 3 |
| chr6:41198325:C:A | missense_variant | TREML2 | 3 |
| chr7:8218358:C:T | missense_variant | ICA1 | 2 |
| chr7:27832847:A:G | missense_variant | JAZF1 | 2 |
| chr7:27895367:T:A | missense_variant | JAZF1 | 4 |
| chr7:27895393:C:T | missense_variant | JAZF1 | 9 |
| chr7:37916516:C:G | missense_variant | SFRP4 | 1 |
| chr7:100319614:A:T | missense_variant | SPDYE3 | 8 |
| chr8:11847759:G:A | missense_variant | CTSB | 1 |
| chr9:104792796:T:G | missense_variant | ABCA1 | 2 |
| chr9:104806324:G:A | missense_variant | ABCA1 | 1 |
| chr9:104816339:G:A | missense_variant | ABCA1 | 1 |
| chr9:104832597:G:A | missense_variant | ABCA1 | 2 |

All families where the varian

| Total Affected Carriers subjects | Total Unaffected Carriers subjects | Mean age-at-onset affected carriers | Mean age-at-onset unaffected carriers | Total families |
|---|---|---|---|---|
| 6 | 0 | 79 | NaN | 4 |
| 4 | 1 | 81.5 | 64 | 1 |
| 5 | 2 | 72.2 | 64.5 | 2 |
| 11 | 4 | 72.90909091 | 64.25 | 6 |
| 6 | 2 | 73.16666667 | 62.5 | 3 |
| 2 | 6 | 81.5 | 71.16666667 | 4 |
| 2 | 1 | 83 | 66 | 2 |
| 3 | 1 | 77 | 71 | 1 |
| 1 | 0 | 67 | NaN | 1 |
| 1 | 0 | 84 | NaN | 1 |
| 1 | 0 | 84 | NaN | 1 |
| 1 | 0 | 74 | NaN | 1 |
| 1 | 0 | 71 | NaN | 1 |
| 1 | 0 | 78 | NaN | 1 |
| 1 | 0 | 87 | NaN | 1 |
| 1 | 2 | 93 | 58 | 1 |
| 1 | 0 | 80 | NaN | 1 |
| 3 | 0 | 71 | NaN | 1 |
| 1 | 0 | 73 | NaN | 1 |

All families where the varian

| Total Affected Carriers subjects | Total Unaffected Carriers subjects | Mean age-at-onset affected carriers | Mean age-at-onset unaffected carriers | Total families |
|---|---|---|---|---|
| 4 | 3 | 83.5 | 70.66666667 | 1 |
| 4 | 1 | 78 | 68 | 3 |
| 1 | 0 | 69 | NaN | 1 |
| 1 | 0 | 60 | NaN | 1 |
| 1 | 0 | 69 | NaN | 1 |
| 7 | 5 | 74.2 | 66.5 | 4 |
| 1 | 0 | 55 | NaN | 1 |
| 1 | 0 | 66 | NaN | 1 |
| 3 | 1 | 85.66666667 | 80 | 1 |

| | | | | |
|---|---|---|---|---|
| 1 | 0 | 85 | NaN | 1 |
| 1 | 0 | 85 | NaN | 1 |

**All families where the varian**

| Total Affected Carriers subjects | Total Unaffected Carriers subjects | Mean age-at-onset affected carriers | Mean age-at-onset unaffected carriers | Total families |
|---|---|---|---|---|
| 4 | 3 | 79.5 | 63.66666667 | 5 |
| 4 | 1 | 72 | 65 | 1 |
| 4 | 0 | 66.5 | NaN | 1 |
| 5 | 0 | 75.6 | NaN | 3 |
| 4 | 0 | 75.75 | NaN | 3 |
| 6 | 2 | 74 | 64.5 | 3 |
| 7 | 4 | 73 | 58.5 | 7 |
| 7 | 2 | 72.57142857 | 64 | 3 |
| 5 | 4 | 67.4 | 56.25 | 1 |
| 4 | 2 | 74.75 | 64.5 | 1 |
| 7 | 4 | 78.28571429 | 65.5 | 3 |
| 8 | 3 | 68.25 | 62.33333333 | 3 |
| 4 | 0 | 83.5 | NaN | 2 |
| 5 | 3 | 67 | 59 | 2 |
| 4 | 0 | 72 | NaN | 3 |
| 12 | 7 | 73.33333333 | 60 | 10 |
| 5 | 2 | 72 | 62 | 3 |
| 4 | 1 | 73.75 | 62 | 2 |
| 5 | 1 | 73.2 | 59 | 3 |
| 4 | 0 | 67.5 | NaN | 3 |
| 4 | 1 | 80.75 | 70 | 4 |
| 4 | 0 | 69.75 | NaN | 1 |
| 4 | 0 | 60.25 | NaN | 3 |
| 7 | 5 | 77 | 71.6 | 8 |
| 10 | 4 | 76.1 | 68 | 7 |
| 1 | 0 | 81 | NaN | 1 |
| 1 | 1 | 83 | 70.75 | 1 |
| 1 | 0 | 78 | NaN | 1 |
| 1 | 0 | 63 | NaN | 1 |
| 1 | 0 | 72 | NaN | 1 |
| 1 | 0 | 76 | NaN | 1 |
| 1 | 1 | 72 | 61 | 1 |
| 1 | 1 | 82 | 63 | 2 |
| 1 | 0 | 67 | NaN | 1 |
| 1 | 0 | 71 | NaN | 1 |

| | | | | |
|---|---|---|---|---|
| 1 | 1 | 72 | 65 | 1 |
| 1 | 1 | 68 | 53 | 1 |
| 1 | 0 | 91 | NaN | 1 |
| 1 | 0 | 74 | NaN | 1 |
| 3 | 2 | 74.83333333 | 68.4 | 3 |
| 1 | 2 | 71.75 | 66 | 2 |
| 4 | 5 | 75.6 | 59 | 8 |
| 1 | 0 | 61 | NaN | 1 |
| 3 | 1 | 73.2 | 57 | 3 |
| 2 | 2 | 78 | 62.25 | 2 |
| 4 | 2 | 72.5 | 60.57142857 | 4 |
| 1 | 1 | 85 | 74 | 1 |
| 1 | 1 | 86 | 62.5 | 1 |
| 1 | 1 | 82.5 | 64 | 1 |
| 2 | 2 | 75 | 63.66666667 | 3 |
| 3 | 3 | 80.5 | 67.5 | 6 |
| 1 | 0 | 85 | NaN | 1 |
| 4 | 2 | 68.55555556 | 62 | 4 |
| 1 | 0 | 80 | NaN | 1 |
| 1 | 0 | 80 | NaN | 1 |
| 1 | 0 | 64 | NaN | 1 |
| 6 | 1 | 72.22222222 | 67 | 6 |
| 1 | 0 | 81 | NaN | 1 |
| 1 | 0 | 90 | NaN | 1 |
| 1 | 0 | 69 | NaN | 1 |
| 1 | 0 | 60 | NaN | 1 |
| 1 | 1 | 86 | 61 | 2 |
| 1 | 2 | 81.5 | 58.5 | 2 |
| 7 | 5 | 73.92857143 | 68.6 | 8 |
| 1 | 1 | 84 | 69 | 1 |
| 1 | 1 | 76.5 | 65.5 | 1 |
| 1 | 0 | 73 | NaN | 1 |
| 1 | 1 | 75.5 | 57 | 1 |
| 1 | 0 | 60 | NaN | 1 |
| 1 | 1 | 75.5 | 68.5 | 1 |
| 1 | 0 | 80 | NaN | 1 |
| 2 | 0 | 64.33333333 | NaN | 2 |
| 4 | 0 | 75.3 | NaN | 4 |
| 1 | 0 | 57 | NaN | 1 |
| 1 | 1 | 75 | 57 | 1 |
| 1 | 1 | 75 | 57 | 1 |
| 1 | 0 | 77 | NaN | 1 |
| 1 | 0 | 71 | NaN | 1 |
| 6 | 3 | 73.75 | 65.33333333 | 8 |
| 1 | 1 | 74 | 61 | 2 |
| 5 | 5 | 80.66666667 | 67.42857143 | 6 |
| 2 | 0 | 55 | NaN | 2 |

| | | | | |
|---|---|---|---|---|
| 2 | 1 | 71 | 64 | 3 |
| 1 | 0 | 77 | NaN | 1 |
| 1 | 1 | 82 | 69 | 1 |
| 7 | 5 | 68.5 | 62.6 | 8 |
| 1 | 0 | 35 | NaN | 1 |
| 1 | 0 | 83 | NaN | 1 |
| 1 | 1 | 78 | 66.5 | 1 |
| 1 | 0 | 59.66666667 | NaN | 1 |
| 1 | 0 | 80 | NaN | 1 |
| 1 | 0 | 65 | NaN | 1 |
| 1 | 0 | 69.33333333 | NaN | 1 |
| 2 | 1 | 78.5 | 66 | 2 |
| 1 | 0 | 74 | NaN | 1 |
| 1 | 1 | 73 | 53 | 1 |
| 1 | 0 | 71 | NaN | 1 |
| 1 | 0 | 64 | NaN | 1 |
| 1 | 0 | 83 | NaN | 1 |
| 1 | 0 | 84 | NaN | 1 |
| 2 | 1 | 75 | 64 | 2 |
| 2 | 2 | 64.25 | 55 | 3 |
| 1 | 0 | 95 | NaN | 1 |
| 2 | 0 | 73.5 | NaN | 2 |
| 1 | 0 | 77 | NaN | 1 |
| 1 | 0 | 66 | NaN | 1 |
| 1 | 1 | 72 | 64.5 | 1 |
| 1 | 0 | 80 | NaN | 1 |
| 1 | 0 | 42 | NaN | 1 |
| 1 | 1 | 77 | 62 | 1 |
| 1 | 0 | 65.25 | NaN | 1 |
| 1 | 0 | 59 | NaN | 1 |
| 3 | 1 | 77.25 | 67 | 3 |
| 2 | 1 | 79.25 | 70 | 2 |
| 1 | 1 | 73 | 62 | 1 |
| 4 | 3 | 75.125 | 57.5 | 5 |
| 1 | 0 | 60 | NaN | 1 |
| 1 | 1 | 77 | 63 | 1 |
| 1 | 0 | 57 | NaN | 1 |
| 1 | 0 | 61 | NaN | 1 |
| 1 | 1 | 75 | 69 | 1 |
| 3 | 2 | 75.2 | 65 | 4 |
| 2 | 4 | 66 | 58.5 | 5 |
| 2 | 0 | 56 | NaN | 2 |
| 1 | 2 | 75.33333333 | 61.5 | 2 |
| 1 | 0 | 72.5 | NaN | 1 |
| 3 | 0 | 67 | NaN | 3 |
| 1 | 0 | 74 | NaN | 1 |
| 11 | 9 | 73.9375 | 65.2 | 17 |

| | | | | |
|---|---|---|---|---|
| 1 | 0 | 72.5 | NaN | 1 |
| 1 | 0 | 65 | NaN | 1 |
| 1 | 1 | 80 | 66 | 1 |
| 1 | 0 | 56 | NaN | 1 |
| 1 | 0 | 70 | NaN | 1 |
| 1 | 0 | 65 | NaN | 1 |
| 1 | 0 | 79 | NaN | 1 |
| 2 | 1 | 64.5 | 59 | 2 |
| 1 | 0 | 35 | NaN | 1 |
| 1 | 0 | 77 | NaN | 1 |
| 1 | 0 | 77 | NaN | 1 |
| 1 | 0 | 52 | NaN | 1 |
| 1 | 1 | 78.5 | 73 | 1 |
| 2 | 2 | 69.66666667 | 62 | 3 |
| 1 | 0 | 67 | NaN | 1 |
| 1 | 1 | 78 | 62 | 1 |
| 1 | 1 | 82.25 | 67 | 1 |
| 1 | 0 | 72 | NaN | 1 |
| 1 | 1 | 72 | 59 | 1 |
| 1 | 0 | 77 | NaN | 1 |
| 2 | 1 | 73 | 63 | 3 |
| 1 | 0 | 75 | NaN | 1 |

**All families where the varian**

| Total Affected Carriers subjects | Total Unaffected Carriers subjects | Mean age-at-onset affected carriers | Mean age-at-onset unaffected carriers | Total families |
|---|---|---|---|---|
| 4 | 1 | 69.5 | 61 | 3 |
| 5 | 4 | 81.6 | 65 | 2 |
| 4 | 0 | 84.25 | NaN | 1 |
| 4 | 3 | 84.25 | 63 | 2 |
| 6 | 5 | 80.66666667 | 64.75 | 3 |
| 5 | 3 | 72.8 | 63.33333333 | 4 |
| 5 | 4 | 73.6 | 62.5 | 4 |
| 7 | 3 | 81.5 | 69.66666667 | 4 |
| 4 | 0 | 79.25 | NaN | 3 |
| 4 | 2 | 62 | 54 | 1 |
| 1 | 0 | 80 | NaN | 1 |
| 1 | 0 | 69 | NaN | 1 |
| 1 | 0 | 68 | NaN | 1 |
| 3 | 0 | 67 | NaN | 1 |
| 1 | 0 | 81 | NaN | 1 |

| | | | | |
|---|---|---|---|---|
| 1 | 0 | 78 | NaN | 1 |
| 1 | 0 | 73 | NaN | 1 |
| 1 | 0 | 73 | NaN | 1 |
| 1 | 0 | 83 | NaN | 1 |
| 1 | 0 | 70 | NaN | 1 |
| 1 | 1 | 81 | NaN | 1 |
| 1 | 0 | 69 | NaN | 1 |
| 2 | 0 | 93 | NaN | 2 |
| 3 | 2 | 80.66666667 | 63 | 1 |
| 1 | 0 | 90 | NaN | 1 |
| 2 | 4 | 72 | 60 | 5 |
| 1 | 0 | 83 | NaN | 1 |
| 1 | 1 | 80 | 60 | 1 |
| 1 | 1 | 89 | 82 | 1 |
| 1 | 0 | 71 | NaN | 1 |
| 1 | 0 | 60 | NaN | 1 |
| 1 | 1 | 84 | NaN | 1 |
| 3 | 5 | 87 | 79.5 | 1 |
| 1 | 0 | 90 | NaN | 1 |
| 2 | 0 | 74.5 | NaN | 1 |
| 2 | 1 | 74.5 | 64 | 1 |
| 1 | 0 | 77 | NaN | 1 |
| 2 | 1 | 72.5 | NaN | 2 |
| 1 | 0 | 72 | NaN | 1 |
| 1 | 0 | 74 | NaN | 1 |
| 1 | 1 | 95 | 88 | 1 |
| 1 | 0 | 62 | NaN | 1 |
| 1 | 0 | 83 | NaN | 1 |
| 1 | 0 | 79 | NaN | 1 |
| 2 | 2 | 88.5 | 77 | 1 |
| 1 | 1 | 89 | 65 | 1 |
| 1 | 0 | 79 | NaN | 1 |
| 1 | 0 | 75 | NaN | 1 |
| 1 | 1 | 84 | 78 | 1 |
| 1 | 0 | 80 | NaN | 1 |
| 2 | 1 | 75.5 | 56 | 2 |
| 3 | 1 | 77.33333333 | NaN | 1 |
| 1 | 2 | 71 | NaN | 1 |
| 1 | 0 | 80 | NaN | 1 |
| 3 | 3 | 72.66666667 | 67 | 4 |
| 1 | 0 | 90 | NaN | 1 |
| 1 | 0 | 81 | NaN | 1 |
| 1 | 0 | 68 | NaN | 1 |
| 1 | 2 | 79 | 69 | 1 |
| 1 | 3 | 82 | 72.33333333 | 1 |
| 1 | 0 | 84 | NaN | 1 |
| 1 | 0 | 69 | NaN | 1 |

| | | | | |
|---|---|---|---|---|
| 1 | 0 | 72 | NaN | 1 |
| 2 | 1 | 77 | 64 | 1 |
| 1 | 0 | 65 | NaN | 1 |
| 1 | 0 | 70 | NaN | 1 |
| 1 | 0 | 70 | NaN | 1 |
| 3 | 4 | 82.66666667 | 68.25 | 1 |
| 1 | 0 | 71 | NaN | 1 |
| 2 | 0 | 63 | NaN | 1 |
| 1 | 0 | 75 | NaN | 1 |
| 2 | 0 | 66 | NaN | 1 |
| 1 | 0 | 72 | NaN | 1 |
| 1 | 0 | 60 | NaN | 1 |
| 1 | 0 | 61 | NaN | 1 |
| 1 | 0 | 86 | NaN | 1 |
| 1 | 0 | 74 | NaN | 1 |
| 1 | 0 | 82 | NaN | 1 |
| 3 | 6 | 80.33333333 | 71.16666667 | 1 |
| 1 | 1 | 65 | NaN | 1 |
| 1 | 0 | 60 | NaN | 1 |
| 2 | 6 | 80.5 | 64.2 | 4 |
| 1 | 0 | 66 | NaN | 1 |
| 1 | 0 | 77 | NaN | 1 |
| 2 | 0 | 74 | NaN | 1 |
| 1 | 0 | 60 | NaN | 1 |
| 1 | 0 | 80 | NaN | 1 |
| 1 | 1 | 68 | NaN | 1 |
| 1 | 1 | 81 | 49 | 1 |
| 1 | 0 | 86 | NaN | 1 |
| 1 | 0 | 90 | NaN | 1 |
| 2 | 2 | 71 | 63.5 | 2 |
| 3 | 0 | 69 | NaN | 2 |
| 3 | 0 | 74 | NaN | 2 |
| 2 | 0 | 74 | NaN | 1 |
| 2 | 0 | 76 | NaN | 1 |
| 1 | 3 | 80 | 67 | 1 |
| 3 | 6 | 80.33333333 | 71 | 1 |
| 1 | 0 | 79 | NaN | 1 |
| 4 | 4 | 85 | 70.75 | 4 |
| 1 | 0 | 67 | NaN | 1 |
| 1 | 1 | 70 | 38 | 2 |
| 1 | 0 | 89 | NaN | 1 |
| 1 | 0 | 66 | NaN | 1 |
| 2 | 0 | 80 | NaN | 1 |

| t is observed | | | |
|---|---|---|---|
| Co-segregation of APOE in all families where the variant is observed (ε4 only;ε3 only;APOE mix) | Number of Families Carrying the variant | Family IDs of families carrying the variant | Segregation type |
| 0;0;4 | 4 | 1177\|1279\|308\|3752 | Incomplete |
| 0;0;1 | 1 | 3761 | Incomplete |
| 0;0;2 | 2 | 303\|3760 | Incomplete |
| 0;0;6 | 6 | 7\|308\|3752\|430\|627 | Incomplete |
| 0;0;3 | 3 | 1563\|1584\|3761 | Incomplete |
| 0;0;4 | 2 | 3761\|858 | Incomplete |
| 0;0;2 | 1 | 1966 | Incomplete |
| 1;0;0 | 1 | 333 | Incomplete |
| 0;0;1 | 1 | 764 | Incomplete |
| 0;1;0 | 1 | 3824 | Incomplete |
| 0;1;0 | 1 | 3824 | Incomplete |
| 0;0;1 | 1 | 3823 | Incomplete |
| 0;0;1 | 1 | 770 | Incomplete |
| 0;0;1 | 1 | 277 | Incomplete |
| 0;0;1 | 1 | 285 | Incomplete |
| 0;0;1 | 1 | 3762 | Incomplete |
| 0;0;1 | 1 | 1917 | Incomplete |
| 0;0;1 | 1 | 769 | Incomplete |
| 0;0;1 | 1 | 793 | Incomplete |

| t is observed | | | |
|---|---|---|---|
| Co-segregation of APOE in all families where the variant is observed (ε4 only;ε3 only;APOE mix) | Number of Families Carrying the variant | Family IDs of families carrying the variant | Segregation type |
| 0;0;1 | 1 | 4_515 | Incomplete |
| 0;0;3 | 2 | 25_41\|4_680 | Incomplete |
| 0;1;0 | 1 | 26_BPC | Incomplete |
| 0;0;1 | 1 | 28_10 | Incomplete |
| 1;0;0 | 1 | 25_76 | Incomplete |
| 2;0;2 | 3 | _128\|10R_R111\|10R_ | Incomplete |
| 0;0;1 | 1 | 11_8 | Incomplete |
| 0;0;1 | 1 | 4_499 | Incomplete |
| 0;0;1 | 1 | 10R_R77 | Incomplete |

| | | | |
|---|---|---|---|
| 0;0;1 | 1 | 25_5 | Incomplete |
| 0;0;1 | 1 | 25_5 | Incomplete |

| t is observed | | | |
|---|---|---|---|
| **Co-segregation of APOE in all families where the variant is observed (ε4 only;ε3 only;APOE mix)** | **Number of Families Carrying the variant** | **Family IDs of families carrying the variant** | **Segregation type** |
| 0;0;5 | 3 | 1177\|164\|308 | Incomplete |
| 0;0;1 | 1 | 317 | Incomplete |
| 0;0;1 | 1 | 3761 | Incomplete |
| 0;1;2 | 3 | 269\|3761\|504 | Incomplete |
| 0;0;3 | 3 | 1177\|212\|3319 | Incomplete |
| 0;0;3 | 2 | 3746\|3823 | Incomplete |
| 2;1;4 | 6 | 7\|1252\|277\|463\|692 | Incomplete |
| 1;0;2 | 3 | 1819\|308\|333 | Incomplete |
| 0;0;1 | 1 | 3761 | Incomplete |
| 0;0;1 | 1 | 343 | Incomplete |
| 0;0;3 | 2 | 1563\|669 | Incomplete |
| 1;0;2 | 3 | 343\|408\|764 | Incomplete |
| 1;1;0 | 2 | 1007\|409 | Incomplete |
| 0;0;2 | 2 | 264\|3752 | Incomplete |
| 1;2;0 | 3 | 203\|3824\|3851 | Incomplete |
| 2;3;5 | 9 | 2193\|285\|3761\|424\|5 | Incomplete |
| 0;0;3 | 2 | 2128\|764 | Incomplete |
| 0;0;2 | 2 | 1582\|317 | Incomplete |
| 1;0;2 | 3 | 1438\|278\|621 | Incomplete |
| 0;0;3 | 3 | 1177\|1317\|3761 | Incomplete |
| 0;2;2 | 3 | 1197\|3087\|3751 | Incomplete |
| 0;0;1 | 1 | 1438 | Incomplete |
| 0;0;3 | 3 | 168\|295\|3761 | Incomplete |
| 1;4;3 | 6 | 328\|3318\|3746\|382 | Incomplete |
| 0;1;6 | 6 | 7\|2193\|236\|318\|380 | Incomplete |
| 0;1;0 | 1 | 3744 | Incomplete |
| 0;0;1 | 1 | 834 | Incomplete |
| 0;1;0 | 1 | 827 | Incomplete |
| 0;1;0 | 1 | 3252 | Incomplete |
| 1;0;0 | 1 | 673 | Incomplete |
| 0;0;1 | 1 | 3541 | Incomplete |
| 0;0;1 | 1 | 167 | Incomplete |
| 0;0;2 | 1 | 764 | Incomplete |
| 0;1;0 | 1 | 434 | Incomplete |
| 0;0;1 | 1 | 1197 | Incomplete |

| | | | |
|---|---|---|---|
| 0;1;0 | 1 | 827 | Incomplete |
| 0;0;1 | 1 | 222 | Incomplete |
| 0;1;0 | 1 | 1346 | Incomplete |
| 0;0;1 | 1 | 3707 | Incomplete |
| 0;0;3 | 3 | 277\|540\|764 | Incomplete |
| 1;0;1 | 1 | 333 | Incomplete |
| 1;2;5 | 4 | 3087\|3762\|3845\|846 | Incomplete |
| 0;0;1 | 1 | 1731 | Incomplete |
| 0;1;2 | 3 | 2075\|627\|863 | Incomplete |
| 0;0;2 | 2 | 3762\|767 | Incomplete |
| 1;0;3 | 4 | 372\|3845\|438\|616 | Incomplete |
| 1;0;0 | 1 | 723 | Incomplete |
| 1;0;0 | 1 | 1403 | Incomplete |
| 0;0;1 | 1 | 1769 | Incomplete |
| 0;1;2 | 2 | 2045\|295 | Incomplete |
| 0;2;4 | 3 | 167\|277\|943 | Incomplete |
| 0;0;1 | 1 | 3324 | Incomplete |
| 0;1;3 | 4 | 197\|303\|380\|746 | Incomplete |
| 1;0;0 | 1 | 1697 | Incomplete |
| 1;0;0 | 1 | 1697 | Incomplete |
| 0;1;0 | 1 | 3824 | Incomplete |
| 2;0;4 | 6 | 7\|277\|3823\|477\|687 | Incomplete |
| 0;0;1 | 1 | 627 | Incomplete |
| 0;0;1 | 1 | 1280 | Incomplete |
| 1;0;0 | 1 | 687 | Incomplete |
| 0;0;1 | 1 | 1438 | Incomplete |
| 1;0;1 | 1 | 622 | Incomplete |
| 0;0;2 | 1 | 20 | Incomplete |
| 3;2;3 | 7 | 150\|307\|3745\|421\|74 | Incomplete |
| 0;0;1 | 1 | 540 | Incomplete |
| 0;0;1 | 1 | 528 | Incomplete |
| 0;0;1 | 1 | 793 | Incomplete |
| 0;0;1 | 1 | 627 | Incomplete |
| 0;0;1 | 1 | 1438 | Incomplete |
| 0;0;1 | 1 | 622 | Incomplete |
| 0;1;0 | 1 | 4 | Incomplete |
| 0;0;2 | 2 | 236\|3761 | Incomplete |
| 0;1;3 | 4 | 1584\|1713\|3761\|746 | Incomplete |
| 0;0;1 | 1 | 361 | Incomplete |
| 0;1;0 | 1 | 380 | Incomplete |
| 0;1;0 | 1 | 380 | Incomplete |
| 0;0;1 | 1 | 285 | Incomplete |
| 0;0;1 | 1 | 277 | Incomplete |
| 1;0;7 | 6 | 7\|167\|222\|317\|372\|3 | Incomplete |
| 0;0;2 | 1 | 1917 | Incomplete |
| 0;1;5 | 5 | 28\|3760\|3761\|665\|69 | Incomplete |
| 0;0;2 | 2 | 277\|765 | Incomplete |

<table>
<tr><td>1;0;2</td><td>2</td><td>1252|342</td><td>Incomplete</td></tr>
<tr><td>1;0;0</td><td>1</td><td>682</td><td>Incomplete</td></tr>
<tr><td>1;0;0</td><td>1</td><td>156</td><td>Incomplete</td></tr>
<tr><td>0;1;7</td><td>7</td><td>[illegible]241|1999|222|285|35[illegible]</td><td>Incomplete</td></tr>
<tr><td>0;0;1</td><td>1</td><td>361</td><td>Incomplete</td></tr>
<tr><td>0;0;1</td><td>1</td><td>317</td><td>Incomplete</td></tr>
<tr><td>0;0;1</td><td>1</td><td>592</td><td>Incomplete</td></tr>
<tr><td>0;0;1</td><td>1</td><td>1317</td><td>Incomplete</td></tr>
<tr><td>0;0;1</td><td>1</td><td>1917</td><td>Incomplete</td></tr>
<tr><td>1;0;0</td><td>1</td><td>446</td><td>Incomplete</td></tr>
<tr><td>0;0;1</td><td>1</td><td>3811</td><td>Incomplete</td></tr>
<tr><td>0;0;2</td><td>2</td><td>1280|2128</td><td>Incomplete</td></tr>
<tr><td>0;0;1</td><td>1</td><td>3707</td><td>Incomplete</td></tr>
<tr><td>0;0;1</td><td>1</td><td>1277</td><td>Incomplete</td></tr>
<tr><td>0;1;0</td><td>1</td><td>319</td><td>Incomplete</td></tr>
<tr><td>0;0;1</td><td>1</td><td>504</td><td>Incomplete</td></tr>
<tr><td>0;0;1</td><td>1</td><td>1280</td><td>Incomplete</td></tr>
<tr><td>0;0;1</td><td>1</td><td>3707</td><td>Incomplete</td></tr>
<tr><td>0;0;2</td><td>2</td><td>1317|528</td><td>Incomplete</td></tr>
<tr><td>0;0;3</td><td>2</td><td>359|764</td><td>Incomplete</td></tr>
<tr><td>0;0;1</td><td>1</td><td>212</td><td>Incomplete</td></tr>
<tr><td>0;0;2</td><td>2</td><td>277|540</td><td>Incomplete</td></tr>
<tr><td>0;0;1</td><td>1</td><td>318</td><td>Incomplete</td></tr>
<tr><td>0;0;1</td><td>1</td><td>3541</td><td>Incomplete</td></tr>
<tr><td>0;0;1</td><td>1</td><td>328</td><td>Incomplete</td></tr>
<tr><td>0;1;0</td><td>1</td><td>3318</td><td>Incomplete</td></tr>
<tr><td>0;0;1</td><td>1</td><td>3324</td><td>Incomplete</td></tr>
<tr><td>0;0;1</td><td>1</td><td>770</td><td>Incomplete</td></tr>
<tr><td>0;0;1</td><td>1</td><td>170</td><td>Incomplete</td></tr>
<tr><td>0;0;1</td><td>1</td><td>1584</td><td>Incomplete</td></tr>
<tr><td>1;1;1</td><td>3</td><td>1403|1928|767</td><td>Incomplete</td></tr>
<tr><td>0;0;2</td><td>2</td><td>3760|477</td><td>Incomplete</td></tr>
<tr><td>0;0;1</td><td>1</td><td>3775</td><td>Incomplete</td></tr>
<tr><td>1;1;3</td><td>4</td><td>[illegible]1279|308|3318|3746[illegible]</td><td>Incomplete</td></tr>
<tr><td>0;1;0</td><td>1</td><td>4</td><td>Incomplete</td></tr>
<tr><td>0;0;1</td><td>1</td><td>770</td><td>Incomplete</td></tr>
<tr><td>0;0;1</td><td>1</td><td>765</td><td>Incomplete</td></tr>
<tr><td>0;0;1</td><td>1</td><td>216</td><td>Incomplete</td></tr>
<tr><td>1;0;0</td><td>1</td><td>1252</td><td>Incomplete</td></tr>
<tr><td>1;1;2</td><td>3</td><td>3823|382|522</td><td>Incomplete</td></tr>
<tr><td>0;1;4</td><td>2</td><td>236|3751</td><td>Incomplete</td></tr>
<tr><td>0;0;2</td><td>2</td><td>3845|859</td><td>Incomplete</td></tr>
<tr><td>0;0;2</td><td>1</td><td>170</td><td>Incomplete</td></tr>
<tr><td>1;0;0</td><td>1</td><td>156</td><td>Incomplete</td></tr>
<tr><td>0;1;2</td><td>3</td><td>257|3761|564</td><td>Incomplete</td></tr>
<tr><td>1;0;0</td><td>1</td><td>156</td><td>Incomplete</td></tr>
<tr><td>3;2;12</td><td>11</td><td>[illegible]303|308|3748|3755|3[illegible]</td><td>Incomplete</td></tr>
</table>

| | | | |
|---|---|---|---|
| 1;0;0 | 1 | 156 | Incomplete |
| 0;0;1 | 1 | 3541 | Incomplete |
| 0;0;1 | 1 | 317 | Incomplete |
| 0;0;1 | 1 | 222 | Incomplete |
| 1;0;0 | 1 | 673 | Incomplete |
| 0;0;1 | 1 | 1317 | Incomplete |
| 1;0;0 | 1 | 409 | Incomplete |
| 0;0;2 | 2 | 360\|421 | Incomplete |
| 0;0;1 | 1 | 361 | Incomplete |
| 0;0;0 | 1 | 4184 | Incomplete |
| 0;0;0 | 1 | 4184 | Incomplete |
| 0;0;1 | 1 | 3762 | Incomplete |
| 1;0;0 | 1 | 3748 | Incomplete |
| 1;1;1 | 2 | 1713\|521 | Incomplete |
| 0;0;1 | 1 | 1317 | Incomplete |
| 0;0;1 | 1 | 958 | Incomplete |
| 0;0;1 | 1 | 372 | Incomplete |
| 0;1;0 | 1 | 211 | Incomplete |
| 0;0;1 | 1 | 167 | Incomplete |
| 1;0;0 | 1 | 3767 | Incomplete |
| 0;1;2 | 2 | 4\|702 | Incomplete |
| 0;0;1 | 1 | 3798 | Incomplete |

| **t is observed** | | | |
|---|---|---|---|
| **Co-segregation of APOE in all families where the variant is observed (ε4 only;ε3 only;APOE mix)** | **Number of Families Carrying the variant** | **Family IDs of families carrying the variant** | **Segregation type** |
| 3;0;0 | 3 | 17_13\|17_25\|4_416 | Incomplete |
| 0;0;2 | 2 | 4_393\|4_557 | Incomplete |
| 0;0;1 | 1 | 4_515 | Incomplete |
| 0;1;1 | 1 | 4_515 | Incomplete |
| 1;1;1 | 3 | 0R_R51\|25_68\|27_13 | Incomplete |
| 0;1;3 | 3 | L0R_R57\|26_EL\|4_55 | Incomplete |
| 2;1;1 | 4 | HOJ\|27_44\|4_589\|4_ | Incomplete |
| 1;2;1 | 4 | 54\|14_1943\|4_606\|8 | Incomplete |
| 2;0;1 | 3 | _HOJ\|27_131\|8_640 | Incomplete |
| 0;0;1 | 1 | 4_553 | Incomplete |
| 0;0;1 | 1 | 8_64037 | Incomplete |
| 0;1;0 | 1 | 26_BPC | Incomplete |
| 0;0;1 | 1 | 19_L0027 | Incomplete |
| 1;0;0 | 1 | 8_64090 | Incomplete |
| 1;0;0 | 1 | 25_68 | Incomplete |

| | | | |
|---|---|---|---|
| 1;0;0 | 1 | 26_SVF | Incomplete |
| 1;0;0 | 1 | 27_247 | Incomplete |
| 1;0;0 | 1 | 27_70 | Incomplete |
| 0;0;1 | 1 | 26_SW | Incomplete |
| 0;0;1 | 1 | 11_8 | Incomplete |
| 0;0;1 | 1 | 26_LWS | Incomplete |
| 0;1;0 | 1 | 26_BPC | Incomplete |
| 0;1;1 | 2 | 4_557\|4_92 | Incomplete |
| 0;1;0 | 1 | 10R_R54 | Incomplete |
| 0;1;0 | 1 | 15_9052 | Incomplete |
| 2;0;3 | 2 | 26_PBO\|4_947 | Incomplete |
| 0;0;1 | 1 | 26_SW | Incomplete |
| 0;1;0 | 1 | 10R_R54 | Incomplete |
| 0;0;1 | 1 | 26_SGA | Incomplete |
| 1;0;0 | 1 | 4_416 | Incomplete |
| 0;0;1 | 1 | 28_10 | Incomplete |
| 1;0;0 | 1 | 10J_123 | Incomplete |
| 0;0;1 | 1 | 4_557 | Incomplete |
| 0;1;0 | 1 | 10R_R54 | Incomplete |
| 0;0;1 | 1 | 10R_R48 | Incomplete |
| 1;0;0 | 1 | 27_247 | Incomplete |
| 0;0;1 | 1 | 27_44 | Incomplete |
| 1;0;1 | 2 | 26_LWS\|4_1836 | Incomplete |
| 0;0;1 | 1 | 4_39 | Incomplete |
| 0;0;1 | 1 | 27_104 | Incomplete |
| 0;0;1 | 1 | 28_12 | Incomplete |
| 0;0;1 | 1 | 27_104 | Incomplete |
| 1;0;0 | 1 | 26_TRL | Incomplete |
| 1;0;0 | 1 | 4_416 | Incomplete |
| 0;0;1 | 1 | 4_515 | Incomplete |
| 0;0;1 | 1 | 2_13 | Incomplete |
| 1;0;0 | 1 | 4_416 | Incomplete |
| 1;0;0 | 1 | 8_64085 | Incomplete |
| 0;0;1 | 1 | 15_10052 | Incomplete |
| 0;0;1 | 1 | 27_134 | Incomplete |
| 2;0;0 | 1 | 8_64090 | Incomplete |
| 0;1;0 | 1 | 22_1 | Incomplete |
| 1;0;0 | 1 | 25_75 | Incomplete |
| 0;0;1 | 1 | 4_680 | Incomplete |
| 1;1;2 | 3 | [illegible]_R51\|15_1121\|19_L[illegible] | Incomplete |
| 0;0;1 | 1 | 4_557 | Incomplete |
| 1;0;0 | 1 | 25_68 | Incomplete |
| 0;0;0 | 1 | 4_3743 | Incomplete |
| 1;0;0 | 1 | 4_1836 | Incomplete |
| 0;0;1 | 1 | 10R_R57 | Incomplete |
| 0;0;1 | 1 | 15_10052 | Incomplete |
| 0;0;1 | 1 | 10J_103 | Incomplete |

| | | | |
|---|---|---|---|
| 1;0;0 | 1 | 27_43 | Incomplete |
| 0;1;0 | 1 | 10R_R54 | Incomplete |
| 1;0;0 | 1 | 25_73 | Incomplete |
| 1;0;0 | 1 | 26_TRL | Incomplete |
| 1;0;0 | 1 | 26_TRL | Incomplete |
| 0;0;1 | 1 | 4_501 | Incomplete |
| 1;0;0 | 1 | 4_364 | Incomplete |
| 0;0;1 | 1 | 11_8 | Incomplete |
| 1;0;0 | 1 | 26_PBO | Incomplete |
| 1;0;0 | 1 | 5_26170 | Incomplete |
| 1;0;0 | 1 | 15_9044 | Incomplete |
| 1;0;0 | 1 | 26_SWM | Incomplete |
| 1;0;0 | 1 | 25_70 | Incomplete |
| 1;0;0 | 1 | 26_CRO | Incomplete |
| 1;0;0 | 1 | 26_HTB | Incomplete |
| 0;0;1 | 1 | 8_64036 | Incomplete |
| 0;0;1 | 1 | 4_501 | Incomplete |
| 1;0;0 | 1 | 25_73 | Incomplete |
| 0;0;1 | 1 | 28_10 | Incomplete |
| 0;1;3 | 1 | 4_393 | Incomplete |
| 1;0;0 | 1 | 8_64045 | Incomplete |
| 1;0;0 | 1 | 8_64085 | Incomplete |
| 1;0;0 | 1 | 10R_R101 | Incomplete |
| 0;1;0 | 1 | 17_17 | Incomplete |
| 1;0;0 | 1 | 25_62 | Incomplete |
| 1;0;0 | 1 | 10J_125 | Incomplete |
| 0;0;1 | 1 | 4_393 | Incomplete |
| 0;0;1 | 1 | 21_ND386 | Incomplete |
| 1;0;0 | 1 | 4_44 | Incomplete |
| 0;0;2 | 2 | 4_39\|8_64036 | Incomplete |
| 0;0;2 | 2 | 26_ANR\|26_QDH | Incomplete |
| 1;0;1 | 2 | 26_HOJ\|4_845 | Incomplete |
| 0;0;1 | 1 | 4_393 | Incomplete |
| 0;0;1 | 1 | 25_57 | Incomplete |
| 0;0;1 | 1 | 4_501 | Incomplete |
| 0;0;1 | 1 | 4_501 | Incomplete |
| 0;0;1 | 1 | 4_499 | Incomplete |
| 0;1;3 | 3 | R_R48\|26_BPC\|8_640 | Incomplete |
| 1;0;0 | 1 | 25_56 | Incomplete |
| 0;0;2 | 1 | 25_61 | Incomplete |
| 0;0;1 | 1 | 4_680 | Incomplete |
| 1;0;0 | 1 | 25_74 | Incomplete |
| 0;0;1 | 1 | 4_393 | Incomplete |
